# Longitudinal multi-platform profiling reveals temporal dynamics of HER2, TROP2, PD-L1 and tumor-infiltrating lymphocytes in triple-negative breast cancer

**DOI:** 10.64898/2026.05.22.26353710

**Authors:** Jorge Gomez Tejeda Zanudo, Busem Binboga Kurt, Allison Frangieh, Alice M. Barkell, John Navarro, Lan Ngo, Ayesha Mohammed-Abreu, Verena M. Prade, Ina Bisha, Sudhanshu Abhishek, Beom-Jun Kim, Melissa Hughes, Karla E. Helvie, Joanna Baginska, David J. Clark, Markus Schick, Ross J. Hill, Tari A. King, Elizabeth A. Mittendorf, Marlon Rebelatto, Eric P. Winer, Sara M. Tolaney, Bruce E. Johnson, Danielle Carroll, Maurizio Scaltriti, Nancy U. Lin, Elza C. de Bruin, Ana C. Garrido-Castro

## Abstract

**Introduction:** With recent approvals of multiple targeted therapies for triple-negative breast cancer (TNBC), including antibody-drug conjugates and immunotherapy in biomarker-selected populations, it is critical to define the temporal evolution of cell-surface target expression from early-stage to metastatic disease, the co-expression patterns across these markers, and optimal quantification methodologies. Here we report biomarker expression profiles measured by multi-omics and pathology-based platforms in patients with TNBC using a large cohort of matched longitudinal tumor samples.

**Methods:** Patients who underwent neoadjuvant chemotherapy (NAC) for stage I-III TNBC or were diagnosed with any stage TNBC and developed metastatic recurrence were retrospectively identified from an institutional database and prospective research metastatic biopsy protocol. Tumor samples from diagnosis (DX), residual disease (RD) post-NAC (if applicable), and metastasis/recurrence (MR) were collected. Quantification of HER2, TROP2, and PD-L1 expression was performed by immunohistochemistry (IHC), whole-exome sequencing, transcriptome sequencing, and targeted mass spectrometry (MS). For HER2, TROP2, and stromal tumor-infiltrating lymphocytes (sTILs), both manual pathologist assessment and computational pathology quantification were obtained. HER2 status was categorized as HER2-0 or HER2-low by local (L-IHC) and central (C-IHC) review, TROP2 status was defined as low (H-score <100), medium (H-score 100-200) or high (H-score >200), and PD-L1 as low (tumor area positivity, TAP <5%) or high (TAP ≥5%). Pathologist-assessed sTILs were classified as low (<10%), medium (≥10% and <40%) or high (≥40%). Biomarkers were compared between primary (DX/RD) and MR, and between pre- vs post-NAC (DX-RD) samples. Correlations between markers, quantification methods, inferred PAM50 subtype, and clinical variables of interest were evaluated.

**Results:** A total of 359 samples from 110 patients with TNBC with data available from at least one platform were included in the analysis. HER2-low prevalence at DX, RD, and MR was: 51% (50/98), 40% (21/53), and 27% (16/60); TROP2 high/medium was 90% (47/52), 91% (42/46), and 88% (28/32); PD-L1-high was 51%, 50%, and 38% (9/24); and sTILs-high/medium was 88% (59/67), 80% (40/50), and 49% (17/35), respectively. While TROP2-high/medium vs low remained stable over time, HER2 IHC and sTILs significantly decreased from DX/RD to MR samples, both at the cohort-level (HER2, p=0.0081; sTILs, p=4.6x10e-5) and longitudinal patient-level (HER2, p=0.030; sTILs, p=0.0077), with a similar decreasing trend for PD-L1 that did not reach statistical significance. HER2 concordance (0 vs low) between L-IHC and C-IHC was 78% (91/116). *ERBB2*, *TACSTD2* and *CD274* mRNA expression were significantly correlated with IHC protein levels, though only *TACSTD2* had limited overlap in distribution of gene expression between high/medium vs low groups. Strong correlation between protein membrane staining intensity from computational pathology, protein expression measured by MS, and pathologist-assed IHC was observed across all biomarkers tested by each method. In comparisons between biomarkers, pathologist-assessed PD-L1 IHC and sTILs were significantly correlated (p=0.0001); 94% (51/54) of PD-L1-high tumors were classified as sTILs high/medium. PAM50 subtype was not significantly correlated with time point or biomarker status, although there was a trend toward more HER2-enriched tumors in HER2-low (20%, 5/25) vs HER2-0 (6%, 3/52) (p=0.086). Across biomarkers and clinical variables, an association between age and sTILs was observed (p=0.038, FDR=0.42) due to a decrease in sTILs high/medium tumors with age, primarily driven by post-treatment (RD/MR) but not DX samples.

**Conclusions:** Multi-platform and multi-omics profiling in this large unique cohort of longitudinal TNBC samples revealed distinct patterns of expression and dynamic changes of key biomarkers of interest for targeted therapies. Given variability with manual IHC scoring, improved methods for quantification of expression may help optimize treatment selection in an individualized manner.

## Introduction

For many years, triple-negative breast cancer (TNBC), defined by the absence of estrogen and progesterone receptor expression and lack of HER2 overexpression or increased copy number, has been considered one of the most challenging breast cancer subtypes to treat, largely due to the lack of effective therapeutic targets, more aggressive tumor biology, and lower survival rates compared to other breast cancer subtypes^1^. Over the past decade, antibody-drug conjugates (ADCs) and immune checkpoint inhibition (ICI) have revolutionized the treatment landscape of early and metastatic TNBC, demonstrating improvements in outcomes compared to more traditional chemotherapy regimens^2–8^.

Currently, there are two ADCs approved by the U.S. Food and Drug Administration (FDA) for the treatment of metastatic TNBC after progression on prior chemotherapy: sacituzumab govitecan (SG, topoisomerase I-inhibitor ADC targeting TROP2^2^) and trastuzumab deruxtecan (T-DXd, topoisomerase I-inhibitor ADC targeting HER2; approved for the approximately 40% of patients with TNBC with HER2-low expression^9^ defined by immunohistochemistry [IHC] staining 1+ or 2+/non-amplified by in situ hybridization^3^). More recently, improvements in outcomes in patients with previously untreated metastatic TNBC treated with SG plus the PD-1 inhibitor pembrolizumab (if PD-L1-positive)^10^ and, if PD-L1-negative, with SG^11^ or datopotamab deruxtecan (topoisomerase I-inhibitor ADC targeting TROP2^12^) were reported compared to physician’s choice of treatment. These TROP2-targeted ADCs are now endorsed by NCCN guidelines as preferred first-line regimens in this setting^13^.

With several approved and emerging ADCs as treatment for breast cancer, including those with overlapping indications and similar target payload classes, it has become imperative to define how to optimally select an initial ADC for a given patient, and if there is a role for sequencing of ADCs. To address these questions, there is a clear need to elucidate the mechanisms that drive response and resistance to ADCs, including how the expression of the ADC target on tumor cells and the surrounding microenvironment may impact the efficacy of the drug. Preliminary data suggest that patients with low levels of TROP2 or HER2 expression still derive benefit from SG or T-DXd, respectively, compared to standard chemotherapy in HER2-negative breast cancer. Thus far, the absolute benefit of these ADCs appears to increase with higher levels of target expression quantified by IHC^14,15^. Importantly, downregulation of target expression has been reported as one of the potential drivers of resistance to ADC therapy. In the DAISY biomarker study of T-DXd, downregulation of HER2 was observed in the majority of patients at the time of progression^15^. Mutations in *TACSTD2* (gene encoding TROP2) that lead to decreased expression of TROP2 on the cell surface membrane and decreased binding of SG to TROP2, have been observed in samples collected after exposure to the TROP2 ADC^16^.

Understanding the role of target expression in the response to ADCs has been further challenged by differences in the assays used and quantification methods. Discordance in HER2 expression measured by IHC has been reported between primary and metastatic tumors in patients with HER2-negative breast cancer defined per ASCO/CAP guidelines, with both gain and loss of expression described^17–19^. Representation of hormone receptor (HR)-negative/HER2-negative (i.e., TNBC) tumors has been limited in these studies, and target expression at multiple time points in TNBC has not been well described. Furthermore, discordance between IHC assays and interobserver variability in pathologist assessment of expression represents a significant challenge in clinical practice^20^. For TROP2, overexpression by IHC has been reported in approximately 85% of TNBC^21^, with no clinical indication for testing to identify patients who may be appropriate candidates for TROP2-directed ADCs. It remains unclear if there is a minimum amount of expression required for these ADCs to be clinically effective. To improve quantification methods of ADC targets, it is critical to understand differences in target expression across assays, including proteomic, transcriptomic and genomic profiling.

Beyond ADCs, pembrolizumab is approved in combination with chemotherapy as first-line therapy for the approximately 40% of patients with metastatic TNBC whose tumor is considered PD-L1-positive (defined in breast cancer by IHC using 22C3 combined positive score [CPS] ≥10^4,5^), and with chemotherapy as neoadjuvant treatment followed by adjuvant therapy for patients with high-risk/early-stage TNBC, unselected by PD-L1 status^6–8^. Emerging data for combination of TROP2 ADCs and ICI in the metastatic setting, including results from the ASCENT-04 trial demonstrating progression-free survival (PFS) benefit with the addition of pembrolizumab to SG as first-line therapy for metastatic PD-L1-positive TNBC^22^, are also changing the treatment algorithm for this biomarker-selected population. In addition, several phase III trials are ongoing exploring TROP2 ADC plus ICI as neoadjuvant or adjuvant therapy for early-stage TNBC (NCT05633654, NCT05629585, NCT06112379, NCT06966700, NCT06393374).

Thus, it is increasingly important to delineate how the expression of cell-surface targets in the tumor and microenvironment evolves over time from early-stage to metastatic breast cancer, the overlap in expression among these targets, and optimal methods to quantify target expression. Here we report the patterns of HER2, TROP2, and PD-L1 expression quantified by different omics and pathology-based platforms, in addition to stromal/intratumoral tumor-infiltrating lymphocytes (sTILs/iTILs), and the overlap and dynamic changes in these markers in patients diagnosed with TNBC using a large collection of matched longitudinal tumor samples.

## Results

### Building an integrated clinical, multi-platform, and multi-omics cohort for longitudinal biomarker assessment in TNBC

An integrated clinical and multi-omics dataset was constructed with comprehensive patient and sample information from medical records, and longitudinal tumor samples for hematoxylin and eosin (H&E) assessment of sTILs and iTILs, quantification by IHC of HER2, TROP2, and PD-L1, and omics (whole-exome sequencing [WES]; transcriptome sequencing, RNA-seq; and targeted mass spectrometry [MS]) data. For HER2, TROP2 and sTILs, both manual and computational pathology quantification was obtained. This dataset was used to assess the dynamics of tumor intrinsic and tumor microenvironment biomarkers of interest from early-stage to metastatic breast cancer, including changes with treatment.

Patients were identified from two sources: 1) an institutional database including all consecutive patients who underwent surgery for stage I-III breast cancer at Dana-Farber Brigham Cancer Center between 2015 (initiation of database) and 2018 (cutoff date for biospecimen requisition), and 2) a prospective research biopsy protocol for patients with metastatic breast cancer. Patients were included in the present analysis if they received neoadjuvant chemotherapy (NAC) for stage I-III TNBC (eTNBC), or if they were diagnosed with any stage TNBC and developed metastatic TNBC (mTNBC). Samples were collected from the following time points: diagnosis (DX), defined as the diagnostic breast biopsy (or surgery, if there was no intervening NAC); residual disease (RD) post-NAC (if applicable); metastasis/recurrence (MR), defined as the first sample collected after unresectable/distant recurrence (after eTNBC) or >30 days after initiation of the first treatment regimen for *de novo* mTNBC. For matched comparisons, if an assay was performed in more than one DX or RD sample, the earliest breast sample was considered; if there was more than one MR sample, the first MR biopsy was considered.

A total of 359 tumor samples (158 DX, 75 RD, 126 MR) from 110 TNBC patients were included in the dataset (**Table 1**, **Fig. 1A-C**). IHC quantification for HER2 (IHC 0, 1+, 2+, and 3+) was abstracted from clinical pathology records (N=308, local IHC [L-IHC]) and reviewed centrally by manual pathologist assessment (N=133, central IHC [C-IHC]). IHC quantification for TROP2 (N=161) and PD-L1 (N=120), and H&E assessment of stromal and intratumoral TILs (N=184, sTILs/iTILs) following the International TILs Working Group guidelines^23^, were performed centrally by manual pathologist assessment (**Fig. 1A**). Computational pathology-based quantification of HER2, TROP2, and sTILs from the digitized slides of the manually scored biomarkers using Quantitative Continuous Scoring (QCS)^24^ was also obtained (**Table 1**).

**Figure 1.**
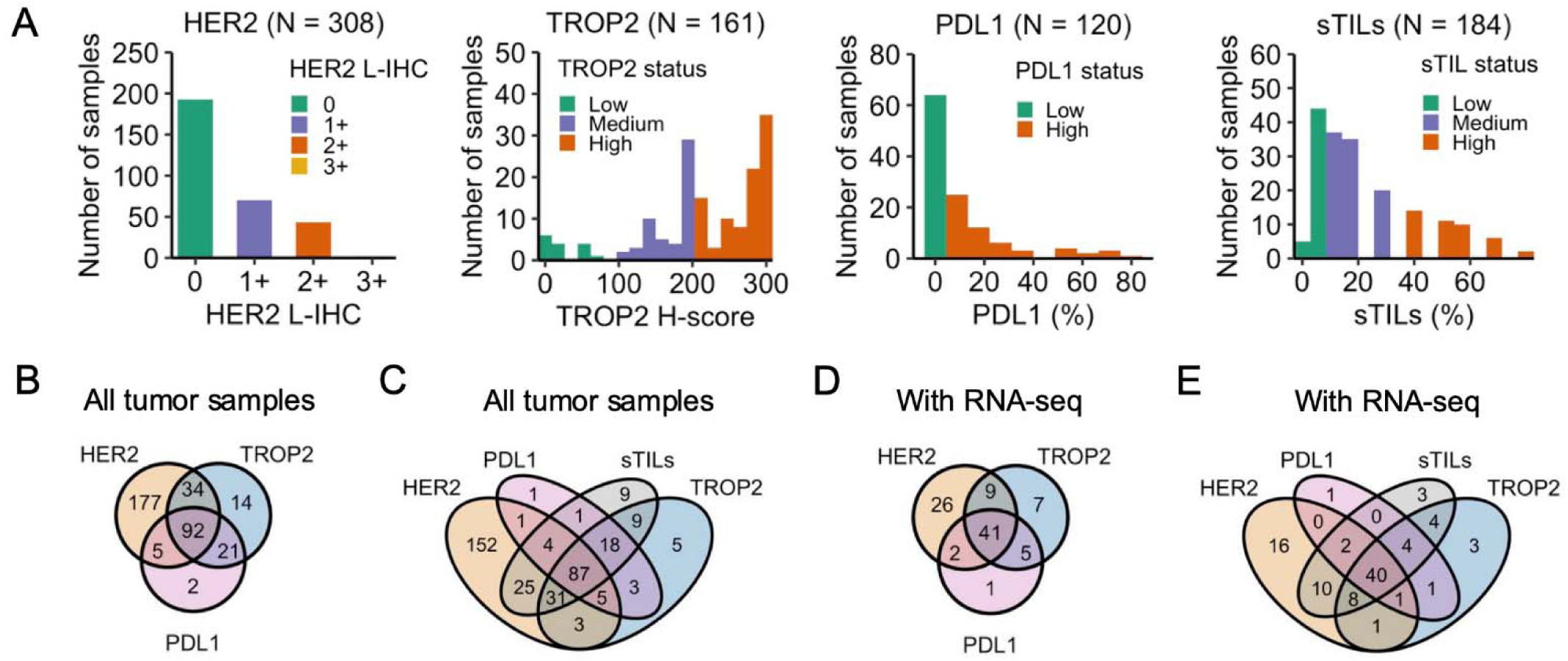
Pathologist-assessed biomarker quantification and status for HER2, TROP2, PD-L1, TILs in TNBC. (A) Distribution of pathologist-assessed biomarker quantification and status. HER2 local IHC (L-IHC) (0, 1+, 2+, 3+); TROP2 IHC H-score: low (H score <100), medium (100-200), high (>200-300); PD-L1 IHC tumor area positivity (TAP): low (TAP <5%), high (TAP ≥5%); stromal TILs (sTILs): low (<10%), sTILs medium (≥10% and <40%), sTILs high (≥40%). (B) Overlap of biomarkers available per sample for HER2 L-IHC, TROP2, and PD-L1 (N=345, 110 pts). (C) Overlap of biomarkers available per sample for HER2 L-IHC, TROP2, PD-L1, and sTILs (N=354, 110 patients). (D) Overlap of biomarkers available per sample with RNA sequencing for HER2 L-IHC, TROP2, and PD-L1 (N=91, 65 pts). (E) Overlap of biomarkers available per sample with RNA sequencing for HER2 L-IHC, TROP2, PD-L1, and sTILs (N=94, 66 patients).

**Table 1.** Patient and sample characteristics of the dataset. Patient and sample characteristics are reported as either median (interquartile range, IQR) or frequency (%).

| **Patient characteristic** | **N = 110 patients** |
| --- | --- |
| Age, median (IQR) | 48 (40-55) |
| Age group |  |
| <40 | 26 (23.6%) |
| 40-60 | 68 (61.8%) |
| >60 | 16 (14.5%) |
| Sex assigned at birth |  |
| Female | 110 (100.0%) |
| Male | 0 (0.0%) |
| Race |  |
| White | 92 (83.6%) |
| Black or African American | 10 (9.1%) |
| Asian | 3 (2.7%) |
| Other | 3 (2.7%) |
| Unknown | 2 (1.8%) |
| Ethnicity |  |
| Hispanic | 7 (6.4%) |
| Non Hispanic | 100 (90.9%) |
| Unknown | 3 (2.7%) |
| Stage at diagnosis |  |
| I | 12 (10.9%) |
| II | 62 (56.3%) |
| III | 21 (19.1%) |
| IV | 10 (9.1%) |
| Unknown<sup>x</sup> | 5 (4.5%) |
| Metastatic disease presentation |  |
| Recurrent | 53 (48.2%) |
| *De novo* | 10 (9.1%) |
| No known metastatic disease at last follow-up | 47 (42.7%) |
| BRCA1/2 germline status |  |
| BRCA1 pathogenic variant | 11 (10.0%) |
| BRCA2 pathogenic variant | 1 (0.9%) |
| BRCA1/2 VUS | 3 (2.7%) |
| BRCA1/2 wild-type | 92 (83.6%) |
| Unknown | 3 (2.7%) |
| Subcohort |  |
| Adj | 20 (18.2%) |
| NAC | 76 (69.1%) |
| dnMBC | 10 (9.1%) |
| Adj/dnMBC<sup>&</sup> | 1 (0.9%) |
| Adj/NAC<sup>&</sup> | 3 (2.7%) |
| **Sample characteristic** | **N = 359 samples** |
| HER2 IHC |  |
| Local | 308 (85.8%) |
| Central (Pathologist-assessed) | 133 (37.0%) |
| Central (Computational pathology) | 85 (23.7%) |
| TROP2 IHC |  |
| Pathologist-assessed | 161 (44.8%) |
| Computational pathology | 144 (40.1%) |
| PDL1 IHC (Pathologist-assessed) | 120 (33.4%) |
| TILs |  |
| sTILs (Pathologist-assessed) | 184 (51.3%) |
| sTILs (Computational pathology) | 104 (29.0%) |
| iTILs (Pathologist-assessed) | 185 (51.5%) |
| Whole-exome sequencing | 84 (23.4%) |
| RNA sequencing | 98 (27.3%) |
| Mass spectrometry | 43 (12.0%) |
| Research-based PAM50* |  |
| Basal | 68 (69.4%) |
| HER2-enriched | 10 (10.2%) |
| Luminal B | 4 (4.1%) |
| Luminal A | 2 (2.0%) |
| Normal | 13 (13.3%) |
| Not classified | 1 (1.0%) |
| Time point |  |
| DX | 158 (44.0%) |
| RD | 75 (20.9%) |
| MR | 126 (35.1%) |
| Receptor subtype (sample) |  |
| TNBC | 244 (68.0%) |
| HR-/HER2+ | 3 (0.8%) |
| HR+/HER2- | 53 (14.8%) |
| HR+/HER2+ | 1 (0.3%) |
| Unknown or indeterminate<sup>§</sup> | 58 (16.2%) |
| Site |  |
| Breast | 211 (58.8%) |
| Axillary Lymph Node | 58 (16.2%) |
| Chest Wall | 24 (6.7%) |
| Liver | 20 (5.6%) |
| Other Lymph Node | 12 (3.3%) |
| Pleural Effusion | 8 (2.2%) |
| Bone | 6 (1.7%) |
| Soft Tissue | 4 (1.1%) |
| Axilla | 4 (1.1%) |
| Lung | 2 (0.6%) |
| Other<sup>#</sup> | 10 (2.8%) |

VUS, variant of unknown significance; IHC, immunohistochemistry; sTILs, stromal tumor-infiltrating lymphocytes; iTILs, intratumoral TILs; DX, diagnosis; RD, residual disease; MR, metastatic/recurrence; TNBC, triple-negative breast cancer; HR, hormone receptor. Subcohorts: Adj, adjuvant (surgery as first intervention for early TNBC); NAC, neoadjuvant (chemotherapy as first intervention for early TNBC); dnMBC (*de novo* metastatic TNBC).
<sup>x</sup>3/5 patients with unknown stage at diagnosis were known to present with early-stage/localized breast cancer, but exact clinical stage at diagnosis was not available.
<sup>&</sup>4 patients had asynchronous primary tumors and were classified in more than one subcohort class.
\*Denominator is the number of samples with available RNA-seq data (n=98).
<sup>§</sup>39 missing all ER/PR/HER2 status information, 13 missing partial ER/PR/HER2 status information (4 HR-/HER2-unknown, 7 HR-unknown/HER2-, 1 HR-unknown/HER2-equivocal, 1 HR-unknown/HER2+), 5 HR-/HER2-equivocal, 1 HR+/HER2-equivocal.
<sup>#</sup>Other sites are abdomen, adrenal, brain, face, jejunum, orbital mass, pericardium, pleura, stomach, and ureter.

In the dataset, HER2, TROP2, and PD-L1 results were available for all three markers in 92 tumor samples (**Fig. 1B**), and a similar number (N=87) had data for all four biomarkers (HER2, TROP2, PD-L1, and TILs) (**Fig. 1C**). The dataset also had quantification of several biomarkers for most tumor samples with omics data. For example, 96% (94/98) of tumor samples with RNA-seq data had at least one type of biomarker data, and 41% (40/98) had data for all four types of biomarkers (HER2, TROP2, PD-L1, and TILs) (**Fig. 1D-E**).

Biomarker data for all four types of biomarkers were available across all time points (**Table 2**, one sample per patient per time point): 211 tumors (98 DX, 53 RD, 60 MR) from 110 patients had HER2 data; 130 tumors (52 DX, 46 RD, 32 MR) from 92 patients had TROP2 data; 105 tumors (41 DX, 40 RD, 24 MR) from 80 patients had PD-L1 data; and 152 tumors (67 DX, 50 RD, 35 MR) from 110 patients had sTILs data. For a subset of these, multiple time points were available: longitudinal DX/RD-MR data for 55, 18, 14, and 23 patients for HER2, TROP2, PD-L1, and sTILs, respectively; longitudinal DX-RD data were available for 46, 20, 10, and 32 patients for HER2, TROP2, PD-L1, and sTILs, respectively.

**Table 2.** Prevalence of biomarker status (pathologist assessment) and PAM50 subtype at different time points in TNBC. One sample per patient for each time point are included. L-IHC, local IHC; DX, at diagnosis; RD, residual disease; MR, metastatic/recurrence.

| **Biomarker status / Sample timepoint** | **DX** | **RD** | **MR** | **Total** |
| --- | --- | --- | --- | --- |
| **HER2 L-IHC (N=211, 110 patients)** |  |  |  |  |
| 0 | 48 (49.0%) | 31 (58.5%) | 44 (73.3%) | 123 (58.3%) |
| 1+ | 34 (34.7%) | 11 (20.8%) | 9 (15.0%) | 54 (25.6%) |
| 2+ | 16 (16.3%) | 10 (18.9%) | 7 (11.7%) | 33 (15.6%) |
| 3+ | 0 (0.0%) | 1 (1.9%) | 0 (0.0%) | 1 (0.5%) |
| Total | 98 (100.0%) | 53 (100.0%) | 60 (100.0%) | 211 (100.0%) |
| **TROP2 status (N=130, 92 patients)** |  |  |  |  |
| Low | 5 (9.6%) | 4 (8.7%) | 4 (12.5%) | 13 (10.0%) |
| Medium | 19 (36.5%) | 17 (37.0%) | 7 (21.9%) | 43 (33.1%) |
| High | 28 (53.8%) | 25 (54.3%) | 21 (65.6%) | 74 (56.9%) |
| Total | 52 (100.0%) | 46 (100.0%) | 32 (100.0%) | 130 (100.0%) |
| **PD-L1 status (N=105, 80 patients)** |  |  |  |  |
| Low | 20 (48.8%) | 20 (50.0%) | 15 (62.5%) | 55 (52.4%) |
| High | 21 (51.2%) | 20 (50.0%) | 9 (37.5%) | 50 (47.6%) |
| Total | 41 (100.0%) | 40 (100.0%) | 24 (100.0%) | 105 (100.0%) |
| **sTILs status (N=152, 97 patients)** |  |  |  |  |
| Low | 8 (11.9%) | 10 (20.0%) | 18 (51.4%) | 36 (23.7%) |
| Medium | 36 (53.7%) | 29 (58.0%) | 11 (31.4%) | 76 (50.0%) |
| High | 23 (34.3%) | 11 (22.0%) | 6 (17.1%) | 40 (26.3%) |
| Total | 67 (100.0%) | 50 (100.0%) | 35 (100.0%) | 152 (100.0%) |
| **PAM50 subtype (N=80, 66 patients)** |  |  |  |  |
| Basal | 20 (76.9%) | 12 (46.2%) | 25 (89.3%) | 57 (71.2%) |
| HER2-enriched | 4 (15.4%) | 1 (3.8%) | 2 (7.1%) | 7 (8.8%) |
| Luminal B | 1 (3.8%) | 1 (3.8%) | 0 (0.0%) | 2 (2.5%) |
| Luminal A | 0 (0.0%) | 0 (0.0%) | 0 (0.0%) | 0 (0.0%) |
| Normal | 0 (0.0%) | 12 (46.2%) | 1 (3.6%) | 13 (16.2%) |
| Not classified | 1 (3.8%) | 0 (0.0%) | 0 (0.0%) | 1 (1.2%) |
| Total | 26 (100.0%) | 26 (100.0%) | 28 (100.0%) | 80 (100.0%) |

When considering one tumor sample per time point per patient, the prevalence of HER2-low tumors (per L-IHC) at DX, RD, and MR, was 51% (50/98), 40% (21/52), and 27% (16/60), respectively (**Table 2**). At these time points, TROP2 high/medium status was 90% (47/52), 91% (42/46), and 88% (28/32), respectively. For PD-L1 IHC, the prevalence of high PD-L1 status was 51% (21/41), 50% (20/40), and 38% (9/24) respectively. High/medium sTILs at DX, RD, and MR was 88% (59/67), 80% (40/50), and 49% (17/35). Most tumors were of basal PAM50 subtype at each time point: 77% (20/26) at DX; 46% (12/26) at RD; and 89% (25/28) at MR.

The availability of multiple biomarker data per sample and across time points allowed us to study the association between biomarkers at the sample-level and their dynamics over time.

### HER2 IHC expression and TILs abundance decreases between primary and metastatic samples at the patient-level and cohort-level

To study the dynamics of biomarkers of interest over time, we compared the frequency of HER2 (HER2-0 vs HER2-low), TROP2 (low vs high/medium), PD-L1 (low vs high) and sTILs (low vs high/medium) between two sets of conditions: (i) primary tumor (DX/RD, at diagnosis or residual disease, if diagnostic tissue was not available) vs metastatic/recurrent (MR) tumor; and (ii) primary tumor (DX) vs residual disease (RD). The comparisons were done at the cohort-level (difference between samples per time point, restricted to one sample per patient per time point) and at the longitudinal patient-level (pairs of samples from the same patient).

Among biomarkers and time point comparisons, whilst TROP2 and PD-L1 status remained stable, there was a statistically significant decrease in HER2 expression by IHC (i.e., decrease of HER2-low, increase of HER2-0) and decrease in TILs abundance (i.e., decrease of sTIL high/medium, increase in sTIL low) between DX/RD and MR samples (**Figs. 2A, 2C**, **Table 2 and 3, Suppl. Tables S1 and S2**). This decrease was seen both at the cohort-level (HER2-0 vs HER2-low, *p* = 0.0081, *OR* = 0.39; sTIL low vs high/medium, *p* = 4.6 × 10^-5^, *OR* = 0.16; two-sided Fisher exact test) (**Fig. 2A**, **Table 2**) and longitudinal patient-level (HER2-0 vs HER2-low, *p* = 0.03, *increase* = 7.3%, *decrease* = 32.7%; sTIL low vs high/medium, *p* = 0.0077, *increase* = 4.3%, *decrease* = 43.5%; two-sided paired Wilcoxon signed rank test) (**Fig. 2C**, **Table 3**).

**Figure 2.**
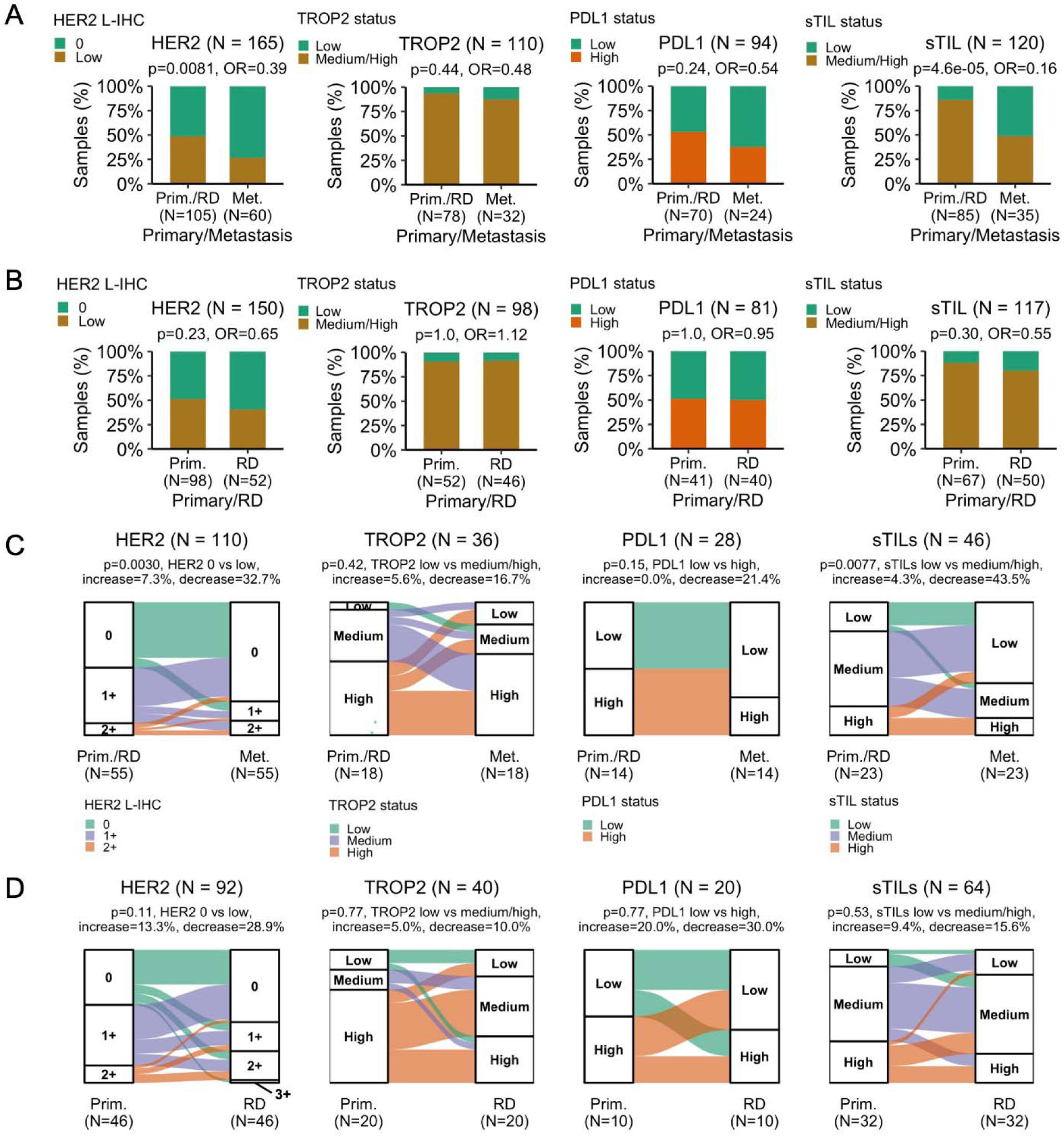
Longitudinal changes in pathologist-assessed biomarkers (HER2, TROP2, PD-L1, TILs) between primary, residual disease, and metastatic samples in TNBC. (A) Biomarkers status between the primary (at diagnosis and/or residual disease post-neoadjuvant therapy) and metastatic setting. One sample per patient for each group/time point are included. *P* values are two-sided Fisher exact test between the shown variables: HER2 L-IHC 0 vs low (1+ or 2+/ISH-), TROP2 low vs medium/high, PD-L1 low vs high, and sTILs low vs medium/high. (B) Biomarkers status between primary (at diagnosis) and residual disease samples. One sample per patient for each group/time point are included. *P* values and comparisons are the same as in Panel A. (C) Biomarkers status between paired primary/metastatic samples from the same patient. *P* values are two-sided paired Wilcoxon signed rank test between: HER2 local IHC (L-IHC) 0 (primary, n=27 pts; metastasis, n=41 pts) vs low (primary, n=28 pts; metastasis, n=14 pts), TROP2 low (primary, n=1; metastasis, n=3) vs medium/high (primary, n=17; metastasis, n=15), PD-L1 low (primary, n=7; metastasis, n=10) vs high (primary, n=7; metastasis, n=4), and sTILs low (primary, n=5; metastasis, n=14) vs medium/high (primary, n=18; metastasis, n=9). (D) Biomarkers status between paired primary/residual disease samples from the same patient. *P* values and comparison are the same as in panel C: HER2 local IHC (L-IHC) 0 (primary, n=18; RD, n=25) vs low (primary, n=27; RD, n=20), TROP2 low (primary, n=3; RD, n=4) vs medium/high (primary, n=17; RD, n=16), PD-L1 low (primary, n=5; RD, n=6) vs high (primary, n=5; RD, n=4), and sTILs low (primary, n=4; RD, n=6) vs medium/high (primary, n=28; RD, n=26).

**Table 3.** Longitudinal changes in biomarker status (pathologist assessment) between different time points in TNBC. L-IHC, local IHC; DX, at diagnosis; RD, residual disease; MR, metastatic/recurrence.

| Biomarker status DX/RD |  | Biomarker status MR |  |  |  |
| --- | --- | --- | --- | --- | --- |
| HER2 L-IHC (N=110, 55 patients) | 0 | 1+ | 2+ | 3+ | Total |
| 0 | 23 (85.2%) | 4 (14.8%) | 0 (0.0%) | 0 (0.0%) | 27 (100.0%) |
| 1+ | 16 (69.6%) | 3 (13.0%) | 4 (17.4%) | 0 (0.0%) | 23 (100.0%) |
| 2+ | 2 (40.0%) | 1 (20.0%) | 2 (40.0%) | 0 (0.0%) | 5 (100.0%) |
| 3+ | 0 (-) | 0 (-) | 0 (-) | 0 (-) | 0 (100.0%) |
| Total | 41 (74.5%) | 8 (14.5%) | 6 (10.9%) | 0 (0.0%) | 55 (100.0%) |
| TROP2 status (N=36, 18 patients) | Low | Medium | High | Total |  |
| Low | 0 (0.0%) | 1 (100.0%) | 0 (0.0%) | 1 (100.0%) |  |
| Medium | 1 (14.3%) | 1 (14.3%) | 5 (71.4%) | 7 (100.0%) |  |
| High | 2 (20.0%) | 2 (20.0%) | 6 (60.0%) | 10 (100.0%) |  |
| Total | 3 (16.7%) | 4 (22.2%) | 11 (61.1%) | 18 (100.0%) |  |
| PD-L1 status (N=28, 14 patients) | Low | High | Total |  |  |
| Low | 7 (100.0%) | 0 (0.0%) | 7 (100.0%) |  |  |
| High | 3 (42.9%) | 4 (57.1%) | 7 (100.0%) |  |  |
| Total | 10 (71.4%) | 4 (28.6%) | 14 (100.0%) |  |  |
| sTILs status (N=46, 23 patients) | Low | Medium | High | Total |  |
| Low | 4 (80.0%) | 1 (20.0%) | 0 (0.0%) | 5 (100.0%) |  |
| Medium | 8 (61.5%) | 5 (38.5%) | 0 (0.0%) | 13 (100.0%) |  |
| High | 2 (40.0%) | 0 (0.0%) | 3 (60.0%) | 5 (100.0%) |  |
| Total | 14 (60.9%) | 6 (26.1%) | 3 (13.0%) | 23 (100.0%) |  |
| PAM50 subtype (N=8, 4 patients) | Basal | non-Basal | Total |  |  |
| Basal | 3 (100.0%) | 0 (0.0%) | 3 (100.0%) |  |  |
| non-Basal | 0 (0.0%) | 1 (100.0%) | 1 (100.0%) |  |  |
| Total | 3 (75.0%) | 1 (25.0%) | 4 (100.0%) |  |  |
| Biomarker status DX |  | Biomarker status RD |  |  |  |
| HER2 L-IHC (N=92, 46 patients) | 0 | 1+ | 2+ | 3+ | Total |
| 0 | 12 (63.2%) | 3 (15.8%) | 3 (15.8%) | 1 (5.3%) | 19 (100.0%) |
| 1+ | 12 (57.1%) | 5 (23.8%) | 4 (19.0%) | 0 (0.0%) | 21 (100.0%) |
| 2+ | 1 (16.7%) | 2 (33.3%) | 3 (50.0%) | 0 (0.0%) | 6 (100.0%) |
| 3+ | 0 (-) | 0 (-) | 0 (-) | 0 (-) | 0 (100.0%) |
| Total | 25 (54.3%) | 10 (21.7%) | 10 (21.7%) | 1 (2.2%) | 46 (100.0%) |
| TROP2 status (N=40, 20 patients) | Low | Medium | High | Total |  |
| Low | 2 (66.7%) | 0 (0.0%) | 1 (33.3%) | 3 (100.0%) |  |
| Medium | 0 (0.0%) | 2 (66.7%) | 1 (33.3%) | 3 (100.0%) |  |
| High | 2 (14.3%) | 7 (50.0%) | 5 (35.7%) | 14 (100.0%) |  |
| Total | 4 (20.0%) | 9 (45.0%) | 7 (35.0%) | 20 (100.0%) |  |
| PDL1 status (N=20, 10 patients) | Low | High | Total |  |  |
| Low | 3 (60.0%) | 2 (40.0%) | 5 (100.0%) |  |  |
| High | 3 (60.0%) | 2 (40.0%) | 5 (100.0%) |  |  |
| Total | 6 (60.0%) | 4 (40.0%) | 10 (100.0%) |  |  |
| sTILs status (N=64, 32 patients) | Low | Medium | High | Total |  |
| Low | 1 (25.0%) | 3 (75.0%) | 0 (0.0%) | 4 (100.0%) |  |
| Medium | 4 (22.2%) | 11 (61.1%) | 3 (16.7%) | 18 (100.0%) |  |
| High | 1 (10.0%) | 5 (50.0%) | 4 (40.0%) | 10 (100.0%) |  |
| Total | 6 (18.8%) | 19 (59.4%) | 7 (21.9%) | 32 (100.0%) |  |
| PAM50 subtype (N=18, 9 patients) | Basal | non-Basal | Total |  |  |
| Basal | 6 (100.0%) | 0 (0.0%) | 6 (100.0%) |  |  |
| non-Basal | 2 (66.7%) | 1 (33.3%) | 3 (100.0%) |  |  |
| Total | 8 (88.9%) | 1 (11.1%) | 9 (100.0%) |  |  |

No statistically significant differences were observed for other biomarkers and time point comparisons (**Figs. 2A-B, 2, Tables 2 and 3**), although there were numerical trends observed both at the cohort-level and longitudinal patient-level. For DX/RD vs MR comparisons, there was a numerical decrease in PD-L1 expression that was seen both at the cohort-level (PD-L1 low vs high, *p* = 0.24, OR = 0.54; two-sided Fisher exact test) (**Fig. 2A**, **Table 2**) and patient-level (PD-L1 low vs high, *p* = 0.15, increase = 0.0%, decrease = 21.4%; two-sided paired Wilcoxon signed rank test) (**Fig. 2C**, **Table 3**). For DX vs RD comparison, there was a similar numerical decrease in HER2 expression that was seen in both groups (HER2-0 vs HER2-low): cohort-level, *p* = 0.23, OR = 0.65 (two-sided Fisher exact test); longitudinal patient-level, *p* = 0.11, increase = 13.3%, decrease = 28.9% (two-sided paired Wilcoxon signed rank test) (**Figs. 2B, 2D**, **Tables 2 and 3**). No statistically significant differences were observed for PAM50 either at the cohort-level (DX vs RD, Basal vs non-Basal, *p* = 0.69, *OR* = 0.56; DX/RD vs MR, Basal vs non-Basal, *p* = 0.17, *OR* = 3.0; two-sided Fisher exact test) or longitudinal patient-level (DX vs RD, Basal vs non-Basal, *p* = 0.48, *Basal to non - Basal* = 0%, *non - Basal to Basal* = 22%; DX/RD vs MR, Basal vs non-Basal, *p* indeterminate, *Basal to non - Basal* = 0%, *non - Basal to Basal* = 0%; McNemar Chi-squared test) (**Tables 2 and 3**).

### Concordance between biomarker quantification methods is highly dependent on both biomarker and testing modality

To characterize the degree of concordance between the different biomarker quantification methods (IHC- and sequencing-based; manual vs computational pathology assessment), we correlated IHC status and/or quantification between these methods for each biomarker of interest (HER2, TROP2, PD-L1, sTILs). We also assessed the correlation between sTILs and iTILs per pathologist-assessment of H&E slides.

For HER2, we compared IHC status abstracted from clinical pathology records (L-IHC) with the central review (C-IHC), the correlation of each with *ERBB2* gene expression from RNA-seq and HER2 protein expression by MS, and their correlation with absolute copy numbers from WES. HER2 L-IHC and C-IHC scores were mostly concordant (N=116 samples, 71 patients), with 72% (N=83/116 samples) matching at the IHC level and 78% (N=91/116 samples) matching by HER2-0 vs HER2-low categories (**Suppl. Table S3**). Most of the discordance observed between HER2-0 and HER2-low (48%, N=12/25) were between C-IHC 1+ and L-IHC 0, reflecting previously described interobserver variability across pathologists^20,25^.

*ERBB2* gene expression was higher in L-IHC HER2-low (N=21) vs HER2-0 (N=46) (*p* = 0.03; Welch’s two-sided t-test), and a similar trend was seen by C-IHC (HER2-low [N=10] vs HER2-0 [N=35], *p* = 0.48; Welch’s two-sided t-test) (**Fig. 3A**). Even though the difference in gene expression between L-IHC HER2-low vs HER2-0 was statistically significant, transcript abundance did not clearly separate the two groups and there was overlap between their distributions (interquartile range [IQR] overlap 29%, L-IHC HER2-0 IQR [0.88-1.12] vs HER2-low IQR [1.05-1.29], normalized *ERBB2* gene expression), which was also observed for C-IHC (IQR overlap 64%, L-IHC HER2-0 IQR [0.86-1.11] vs HER2-low IQR [0.95-1.25], normalized *ERBB2* gene expression). A similar overlap between the distributions of *ERBB2* gene expression was seen in HER2-low vs HER2-0 IHC groups in an independent MBC cohort with all receptor subtypes (The Metastatic Breast Cancer Project^26^), although this overlap was not observed when comparing HER2-positive vs HER2-low or HER2-0 tumors (**Suppl. Fig. S1B**). In contrast to *ERBB2* gene expression, HER2 protein expression, as determined by MS, was not only significantly higher in HER2-low vs HER2-0 (L-ICH *p* = 0.0097; C-IHC *p* = 027; Welch’s two-sided t-test), but there was also similar or less overlap between the distributions of expression (L-IHC, IQR overlap 16%, HER2-0 IQR [197-452] amol/μg vs HER2-low IQR [412-1252] amol/μg, HER2 protein expression by MS; C-IHC, IQR overlap 42%, HER2-0 IQR [318-671] amol/μg vs HER2-low IQR [521-1105] amol/μg, HER2 protein expression by MS) (**Suppl. Fig S2A**). Similarly to HER2 protein expression by MS, HER2 membrane staining intensity (SI) from computational pathology was both significantly higher in HER2-low vs HER2-0 (L-IHC *p* = 0.0061; Welch’s two-sided t-test; C-IHC *p* = 2.1 × 10^-5^; Welch’s two-sided t-test) (**Fig. 3C**, **Suppl. Fig. S3A**), and had similar or less overlap between the distributions of SI compared to *ERBB2* gene expression (L-IHC, IQR overlap 33%, HER2-0 IQR [5.2-7.6] vs HER2-low IQR [6.8-13.2], HER2 membrane SI; C-IHC, IQR overlap 0%, HER2-0 IQR [5.3-7.3] vs HER2-low IQR [8.8-13.9], HER2 membrane SI).

**Figure 3.**
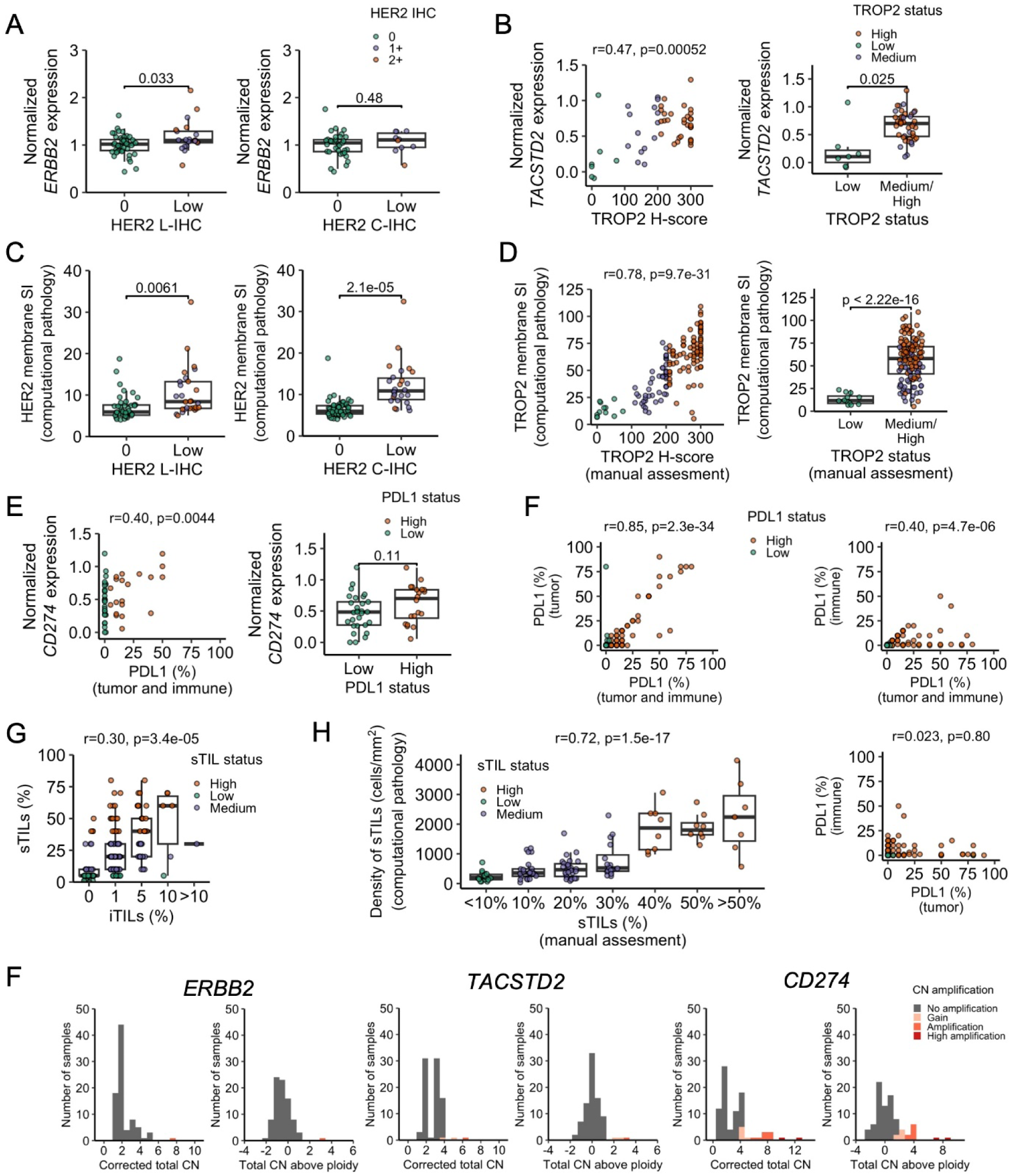
Correlation of biomarker quantification across different platforms in TNBC. (A) Local (L-IHC) and central (C-IHC) HER2 IHC status vs *ERBB2* gene expression from RNA sequencing. HER2-0 L-IHC, N=46 samples; HER2-low L-IHC, N=21 samples; C-IHC HER2-0 N=35 samples; C-IHC HER2-low N=10 samples. (B) TROP2 pathologist-assessed quantification and *TACSTD2* gene expression from RNA sequencing (N=51 samples). TROP2 low, N=7 samples; TROP2 medium/high, N=44 samples. (C) Local (L-IHC) and central (C-IHC) HER2 IHC status from manual pathologist assessment vs HER2 QCS median membrane staining intensity (SI) from computational pathology. HER2-0 L-IHC, N=42 samples; HER2-low L-IHC, N=28 samples; C-IHC HER2-0 N=60 samples; C-IHC HER2-low N=26 samples. (D) TROP2 manual pathologist-assessed quantification vs TROP2 QCS median membrane SI from computational pathology (N=144 samples). TROP2 low, N=12 samples; TROP2 medium/high, N=132 samples. (E) PD-L1 tumor area positivity from tumor and immune cells and *CD274* gene expression from RNA sequencing (N=49 samples). PD-L1 low, N=29 samples; PD-L1 high, N=20 samples. (F) PD-L1 tumor area positivity from tumor cells, immune cells, and both (N=120 samples). (G) Distribution of stromal and intratumoral TILs (N=184 samples). (H) Stromal TILs status by manual pathologist assessment vs stromal TILs density from computational pathology (N=104 samples). (I) Distribution of *ERBB2*, *TACSTD2*, and *CD274* copy numbers (purity/ploidy corrected copy numbers from whole-exome sequencing) (N=84 samples). RNA-seq expression is measured in upper quartile normalized log2(TPM+1). Welch’s t-test (two-sided) is used for statistical comparisons between groups. Pearson correlation (two-sided) is used for statistical correlation between variables.

For TROP2, we compared IHC, both as a continuous (H-score) and categorical (high/medium vs low) variable, with *TACSTD2* gene expression from RNA sequencing data, TROP2 protein expression by MS, and TROP2 membrane SI from computational pathology. *TACSTD2* gene expression was well-correlated with H-score (N=51, Pearson r=0.47, *p* = 0.00052) and was significantly higher in TROP2 high/medium (N=44) vs low (N=7) tumors (*p* = 0.025; Welch’s two-sided t-test) (**Fig. 3B**). In contrast to HER2, there was limited overlap in the distribution of *TACSTD2* gene expression between TROP2 IHC groups (IQR overlap 0%, TROP2 high/medium IQR [0.47-0.82] vs TROP2 low IQR [0-0.22], normalized *TACSTD2* gene expression). A stronger correlation was observed between TROP2 protein expression by MS and IHC H-score (N=43, Pearson r=0.69, *p* = 2.4 × 10^-7^), along with higher TRO2 protein expression by MS in TROP2 IHC high/medium (N=37) vs low (N=6) tumors (*p* = 2.9 × 10^-9^; Welch’s two-sided t-test) (**Suppl. Fig S2B**). Similarly, a strong correlation was observed between TROP2 membrane SI from computational pathology and IHC H-score from manual assessment (N=144, Pearson r=0.78, *p* = 9.7 × 10-^31^), together with higher TROP2 membrane SI in TROP2 IHC high/medium (N=132) vs low (N=12) tumors (*p* < 2.2 × 10^-16^; Welch’s two-sided t-test) (**Fig. 3D, Suppl. Fig. S3B**).

For PD-L1, we first compared TAP (combined tumor/immune cell staining) scoring of IHC stained slides with *CD274* gene expression from bulk RNA-seq, and then compared IHC (%) between tumor, immune, and TAP scores. *CD274* expression was significantly correlated with TAP as a continuous variable (N=49, Pearson r=0.40, *p* = 0.0044) and was numerically higher in PD-L1 IHC high (N=20) vs low (N=29) tumors (*p* = 0.11; Welch’s two-sided t-test) (**Fig. 3E**). Although PD-L1 TAP (%) and gene expression were correlated, this appeared to be primarily driven by concordance in 5 samples with high values for both quantities (**Fig. 3E, left**). Consistent with this observation, the distribution of gene expression for PD-L1 IHC high vs low was overlapping (PD-L1 high IQR [0.39-0.84] vs low IQR [0.28-0.65], normalized *CD274* gene expression) and the range of gene expression for tumors with PD-L1 TAP 0% spanned the same values as those with PD-L1 TAP ≥5% (i.e., high) (**Fig. 3E, left**). Percentage of PD-L1-positivity between tumor cells and immune cells was not correlated (N=120, Pearson r=0.02, p= 0.8), and the combined TAP score was more strongly correlated with that of tumor cells (Pearson r=0.85, *p* = 2.3 × 10^-34^) than that of immune cells (Pearson r=0.40, *p* = 4.7 × 10^-6^) (**Fig. 3F**). In contrast to *CD274* gene expression, PD-L1 protein expression by MS and PD-L1 TAP (%) had a much stronger correlation (N=38, Pearson r=0.96, *p* = 7.3 × 10^-21^) and did not show the full range of MS protein expression for PD-L1 TAP 0%. In addition, PD-L1 protein expression by MS was significantly higher in PD-L1 IHC high (N=15) vs PD-L1 low (N=23) tumors (*p* = 0.042; Welch’s two-sided t-test), and had less overlap in distribution between PD-L1 IHC groups (PD-L1 high IQR [22-67] vs low IQR [3-22], PD-L1 protein expression by MS) **(Suppl. Fig S2C).**

For TILs, we compared sTILs and iTILs (both as continuous and categorical variables), and also manual sTILs assessment with sTILs density derived from computational pathology. A significant correlation was observed between the percentage of sTILs and iTILs (N=184, Pearson r=0.30, *p* = 3.4 × 10^-5^), with the dynamic range of sTILs spanning almost 0-50% while iTILs spanned mostly 0-5% (sTILs IQR [5-30%], 10-90 percentile [5-50%]; iTILs IQR [0-1%], 10-90 percentile [0-5%]) (**Fig. 3G**). The distribution of sTILs by iTILs had a minimal to moderate amount of overlap and this depended on the value (IQR overlap 0%, 50%, 0% respectively for iTILs 0% vs 1%, iTILs 1% vs 5%, and iTILs 0% vs 5%) (iTILs 0% IQR [5-10] vs 1% IQR [10-30] vs 5% IQR [20-50], sTILs %). sTILs and iTILs status were correlated (*p* = 7.8 × 10^-6^; two-sided Fisher exact test), with 98% (N=41/42) of iTILs high tumors being classified as sTILs high/medium, and 34% of iTILs low (N=48/142) being classified as sTILs low (**Suppl. Table S4**). Percentage of sTILs from manual assessment was strongly correlated with sTILs density from computational pathology (N=104, Pearson r=0.72, *p* = 1.5 × 10^-17^) (**Fig. 3H**, **Suppl. Fig. S3C**), and there was a strong separation in sTILs density when comparing sTILs low vs medium vs high (sTILs low vs medium *p* = 1.3 × 10^-5^, sTILs low vs high *p* = 6.8 × 10^-10^, sTILs medium vs high *p* = 2.6 × 10^-8^; Welch’s two-sided t-test) (IQR overlap 0.8%, 0%, 0%, respectively for sTILs low vs medium, low vs high, medium vs high) (sTILs low [136-305] cells/mm^2^, sTILs medium [304-646] cells/mm^2^, sTILs high [1430-2360] cells/mm^2^).

Comparing copy number alterations in *ERBB2*, *TACSTD2*, and *CD274* from WES with IHC protein and gene expression did not yield any statistically significant correlations (Pearson *p* > 0.05). In particular, high amplifications of *ERBB2* were not observed in any tumor samples, as expected in TNBC (**Fig. 3I**), and *ERBB2* copy numbers were not higher in HER2-low compared to HER2-0 samples (**Suppl. Fig. S1A**). Consistent with these data, *ERBB2* copy number was not significantly higher in HER2-low or HER2-0 samples from The Metastatic Breast Cancer Project, but HER2-low samples had numerically higher copy number (*p* = 0.11; Welch’s two-sided t-test) (**Suppl. Fig S1B**). To control for the quality of our copy number data, we verified that amplifications in *MYC*, a frequently amplified gene in TNBC, were common among the samples in our cohort (**Suppl. Fig S1C**). Additionally, we verified that the distribution of both *MYC* and *ERBB2* copy numbers was similar to that described for patients with TNBC from The Metastatic Breast Cancer Project and distinct to the highly amplified *ERBB2* copy number profile in patients with HER2-positive tumors (**Suppl. Fig S1C**).

### PD-L1 positivity correlates with high TILs abundance across timepoints

To evaluate the correlation among pathologist-assessed markers of interest, we performed paired comparisons of biomarker status (HER2, TROP2, PD-L1, sTILs) in tumor samples from the cohort.

Among all comparisons between biomarkers, there were no statistically significant correlations observed for HER2 or TROP2 (*p* > 0.05 two-sided Fisher exact test) (**Fig. 4, Suppl. Table S5**). The only significant correlation was between PD-L1 and sTILs status (PD-L1 vs sTILs, p= 0.00011, FDR = 6.4 × 10^-4^, *OR* = 0.11, two-sided Fisher exact test), with 94% (N=51/54) of PD-L1-high samples categorized as sTILs high/medium, and 36% (N=20/56) of PD-L1-low samples categorized as sTILs low (**Fig. 4C**, **Table 4, Suppl. Table S5**). The correlation between PD-L1 and sTILs status was not driven by samples from a specific time point, given that a similar numerical correlation and trend in significance or statistical significance was seen in DX (*p* = 0.13, two-sided Fisher exact test), RD (*p* = 0.017, two-sided Fisher exact test), and MR (*p* = 0.10, two-sided Fisher exact test) samples (**Table 4**).

**Figure 4.**
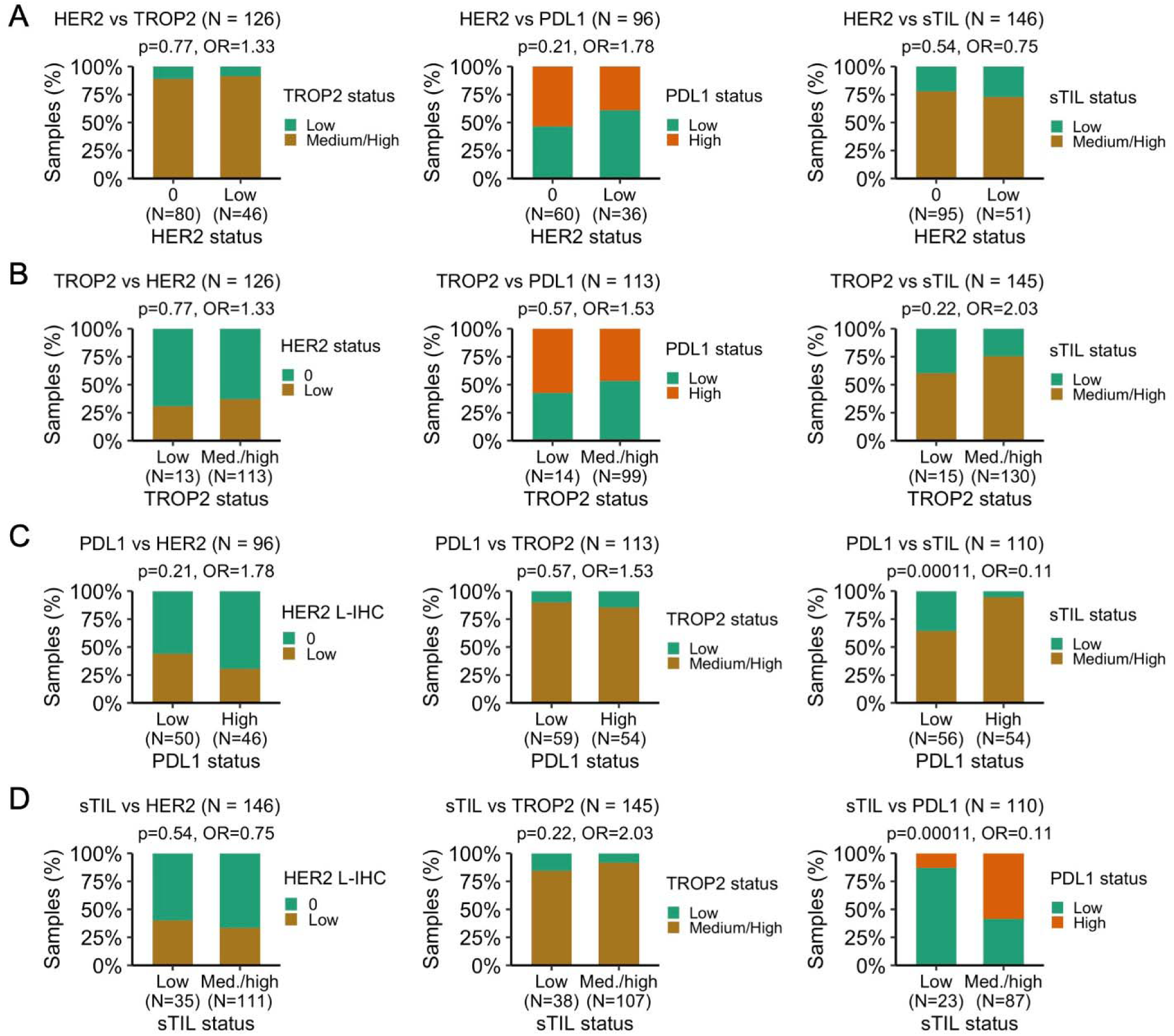
Correlation between pathologist-assessed biomarkers (HER2, TROP2, PD-L1, TILs) in TNBC. (A) Correlation between HER2 local IHC (L-IHC) status (0, low) and other biomarker status (TROP2, PD-L1, and sTILs). (B) Correlation between TROP2 status (low, medium/high) and other biomarker status (HER2, PD-L1, and sTILs). (C) Correlation between PD-L1 status (low, medium) and other biomarker status (HER2, PD-L1, and sTILs) (D) Correlation between sTILs status (low, medium/high) and other biomarker status (HER2, PD-L1, and sTILs) *P* values are two-sided Fisher exact test between the variables in each plot.

**Table 4.** Correlation between PD-L1 status and sTILs status (pathologist assessment) in TNBC. *P* values are a two-sided Fisher exact test between PD-L1 low vs high and sTILs low vs medium/high.

| **PD-L1 status/sTILs status (N=110, 77 patients)** | Low | Medium | High | Total | *P* |
| --- | --- | --- | --- | --- | --- |
| Low | 20 (35.7%) | 33 (58.9%) | 3 (5.4%) | 56 (100.0%) | 1.07E-04 |
| High | 3 (5.6%) | 29 (53.7%) | 22 (40.7%) | 54 (100.0%) |  |
| Total | 23 (20.9%) | 62 (56.4%) | 25 (22.7%) | 110 (100.0%) |  |
| **PD-L1 status/sTILs status DX (N=44, 40 patients)** | Low | Medium | High | Total | *P* |
| Low | 6 (28.6%) | 13 (61.9%) | 2 (9.5%) | 21 (100.0%) | 0.126 |
| High | 2 (8.7%) | 9 (39.1%) | 12 (52.2%) | 23 (100.0%) |  |
| Total | 8 (18.2%) | 22 (50.0%) | 14 (31.8%) | 44 (100.0%) |  |
| **PD-L1 status/sTILs status RD (N=36, 34 patients)** | Low | Medium | High | Total | *P* |
| Low | 4 (28.6%) | 10 (71.4%) | 0 (0.0%) | 14 (100.0%) | 0.017 |
| High | 0 (0.0%) | 15 (68.2%) | 7 (31.8%) | 22 (100.0%) |  |
| Total | 4 (11.1%) | 25 (69.4%) | 7 (19.4%) | 36 (100.0%) |  |
| **PD-L1 status/sTILs status MR (N=30, 23 patients)** | Low | Medium | High | Total | *P* |
| Low | 10 (47.6%) | 10 (47.6%) | 1 (4.8%) | 21 (100.0%) | 0.100 |
| High | 1 (11.1%) | 5 (55.6%) | 3 (33.3%) | 9 (100.0%) |  |
| Total | 11 (36.7%) | 15 (50.0%) | 4 (13.3%) | 30 (100.0%) |  |

### Biomarker status and clinical characteristics beyond primary/metastatic timepoint are not strongly correlated

HER2 status, but not TROP2, PD-L1 and sTILs status, was found to significantly differ between primary and metastatic samples at both the cohort and patient-level. To investigate if other clinical and sample attributes were associated with biomarker status, we evaluated the correlation with a selection of patient-level variables (race, age, stage at initial diagnosis, metastatic presentation [*de novo* vs recurrent], germline *BRCA1/2* status [pathogenic vs wild-type; variants of unknown significance excluded], subcohort based on initial treatment strategy [adjuvant: early TNBC, surgery as first intervention; neoadjuvant: early TNBC, neoadjuvant chemotherapy as first intervention; dnMBC, *de novo* metastatic TNBC]) and sample-level variables (research-based PAM50 subtype, sample site).

For patient-level variables, there were no statistically significant correlations between biomarker status and these variables after correcting for multiple comparisons (FDR < 0.10, two-sided Fisher exact test) (**Suppl. Table S6**). For sample-level variables, PAM50 subtype was not significantly correlated with biomarker status, although there was a trend that did not reach statistical significance for HER2 status driven by a numerical increase of HER2-enriched tumors in the HER2-low (20%, N=5/25) compared to the HER2-0 (6%, N=3/52) group (*p* = 0.086, two-sided Fisher exact test) (**Suppl. Table S7**). Site was significantly associated with HER2 status, with the effect being driven by a high proportion of HER2-low (74%, N=84/113) vs HER2-0 (57%, N=110/193) tumors among breast samples (**Suppl. Table S8**), consistent with the observed decrease in HER2 level from primary to metastatic samples (**Figs. 2A, 2C**).

As an exploratory analysis, we investigated the top biomarker/patient-level comparisons that reached or trended toward statistical significance (p <0.1) prior to multiple comparison correction: sTILs and age (*p* = 0.038, two-sided Fisher exact test), sTILs and subcohort (adjuvant vs neoadjuvant vs dnMBC) (*p* = 0.045, two-sided Fisher exact test), TROP2 and stage (*p* = 0.080, two-sided Fisher exact test), and PD-L1 and age (*p* = 0.099, two-sided Fisher exact test) (**Suppl. Table S6**). The association between sTILs status and subcohort was driven by an increase in sTILs high/medium samples in patients who received neo-adjuvant therapy (82%, N=90/110) vs those in the adjuvant (65%, N=17/26) or dnMBC group (56%, N=5/9), and this trend was only observed in DX samples (**Suppl. Table S9**).

The association between age and sTILs status was due to a decrease in sTILs high/medium with age (<40 years, 83%, N=29/35; 40-60 years, 79%, N=74/94; >60 years, 57%, N=13/23), and this effect was driven by RD and MR, but not DX samples (**Suppl. Table S10**). The numerical association between age and PD-L1 status reflected a decrease in PD-L1 high tumors with age (<40 years, 62%, N=13/21; 40-60 years, 46%, N=31/67; >60 years, 35%, N=6/17) (**Suppl. Table S10**). The numerical association between stage and TROP2 status reflected a reduced proportion of patients with stage III breast cancer in TROP2 low tumors (stage I-II, 92%, N=12/13; stage III, 0%, N=0/13; stage IV, 8%, N=1/13) vs TROP2 high/medium tumors (stage I-II, 65%, N=72/111; stage III, 28%, N=31/111; stage IV, 7%, N=8/111) (**Suppl. Table S9**).

### Case studies of patients treated with HER2-directed and TROP2-directed ADCs

To assess whether baseline expression and changes in HER2 or TROP2 status were associated with response to ADCs targeting these markers, we identified patients exposed to HER2- and/or TROP2-targeted ADCs. 8/110 patients (7.2%) were exposed to an ADC, with 6 patients receiving TROP2 ADCs (1 SG, 4 TROP2 ADCs in a clinical trial, and 1 both SG and a TROP2 ADC in a trial), 1 patient receiving HER2 ADC (T-DM1), and 1 patient receiving both HER2 and TROP2 ADCs (T-DM1, T-DXd, and a TROP2 ADC in a trial). Of these, all had biomarker status of the ADC target either from pathology assessment (7/8) or inferred from RNA-seq data (1/8, TROP2 status inferred from RNA-seq). Three patients had biomarker status both before and after receiving ADC therapy (case 300435, TROP2 ADC; case 300371, T-DM1; case 300511, T-DM1). Note that the low prevalence of ADC exposure can be explained by the fact that, for all patients diagnosed with metastatic breast cancer (N=63 patients, 59 patients with available diagnosis date), the metastatic diagnosis date preceded the approval of the first next-generation ADC for TNBC (SG in May 2020).

For the three patients with biomarker status both before and after receiving an ADC, we found that all exhibited a decrease in their respective biomarker levels (**Fig. 5A-C**). For case 300435, a TROP2-targeted ADC was administered 26 months after dnMBC diagnosis, with a time-on-treatment of 1.5 months, where TROP2 status was medium (IHC, H-score 200, pathologist assessed) in an initial diagnostic sample, and high/medium (inferred from RNA-seq) in a sample collected two months before starting ADC treatment. However, following ADC, TROP2 status was determined to be low (IHC, H-score 25, pathologist assessed) or medium/low (inferred from RNA-seq) from a sample collected 13 months after the ADC (**Fig. 5A**). For cases 300371 and 300511, T-DM1 was administered because of a HER2-positive (300371) or HER2-equivocal sample (300511) in the metastatic setting (31 and 62 months after primary diagnosis) with limited time-on-treatment (300371, 1.5 months; 300511, 3 months). In both cases, the HER2 status prior to the ADC was obtained from the primary (300371, IHC 2+/in situ hybridization [ISH] non-amplified; 300511, IHC 1+) and metastatic (300371, IHC 3+ and IHC 2+/ISH equivocal; 300511 IHC 2+/ISH unknown) diagnosis, and it decreased when compared to those obtained after the ADC (300371, IHC 0 and IHC 1+, 6 and 9 months after the ADC; 300511, IHC 0, 38 months after the ADC). Whilst these data suggest that ADC exposure can drive a reduction in target expression, the receipt of multiple treatments in between samples and the limited time-on-treatment with the ADC makes it difficult to conclusively link the reduction in biomarker expression to ADC exposure.

**Figure 5.**
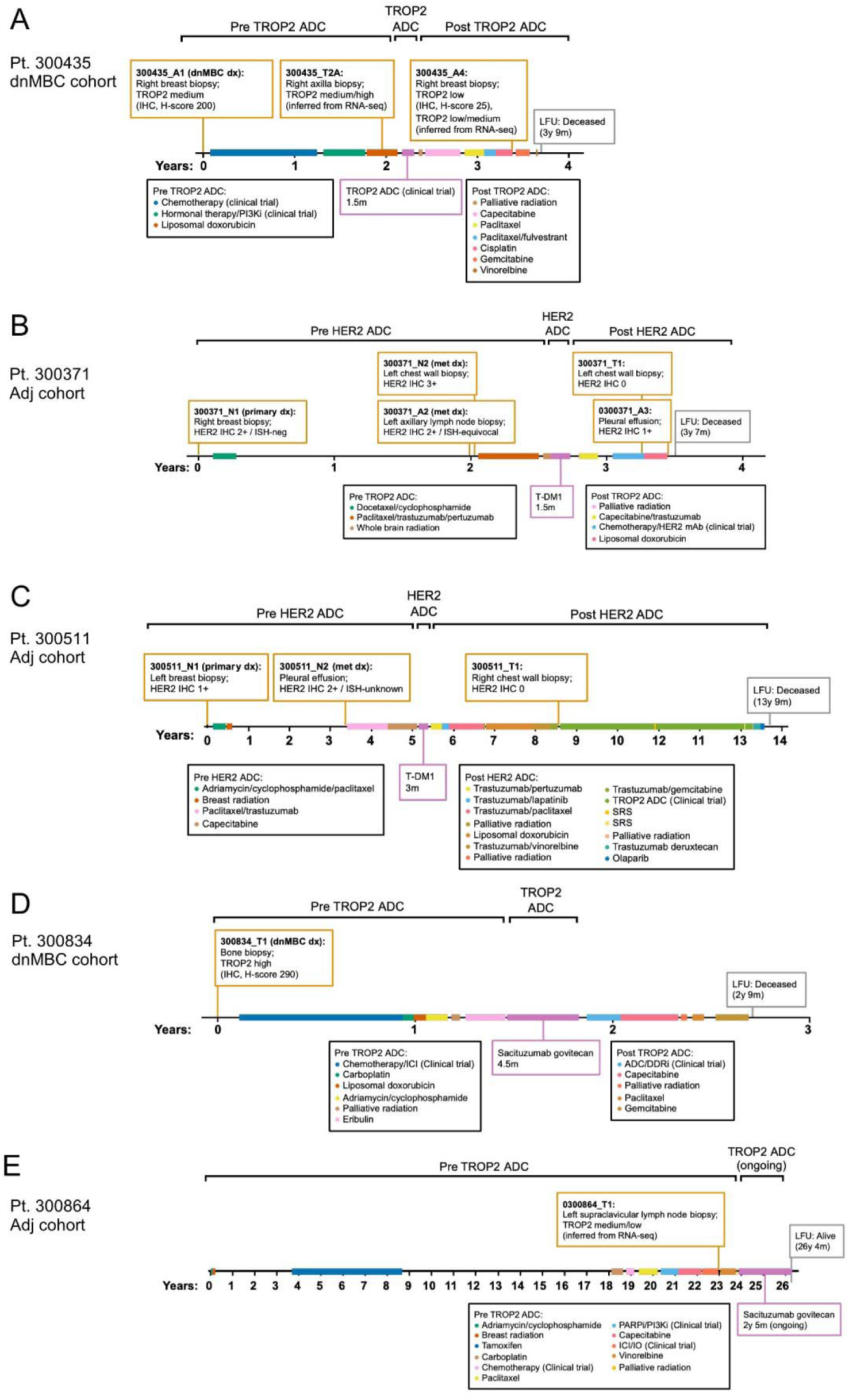
Timelines in patients who received TROP2-directed or HER2-directed antibody-drug conjugates. (A-C) Patients treated with TROP2 (pt. 300345, panel A) or HER2 ADCs (pt. 300371, panel B; pt. 300511, panel C) with biomarker status in the ADC target before and after ADC treatment. (D-E) Selected patients treated with TROP2 ADCs that were TROP2-high and had limited response (pt. 300834, panel D) or were TROP2-low and had a durable response (Pt. 300864, panel E).

For the 7 patients who received TROP2-targeted ADCs (focusing on the first ADC for the patient receiving multiple regimens), there was no clear relationship between the most recent TROP2 status (5/7 IHC, pathologist assessed; 2/7 inferred from RNA-seq) and time-on-treatment with TROP2 ADCs, although TROP2 status was evaluated in samples collected more than 17 months before treatment in most cases (median 18 months prior to treatment start, IQR [7-25]). For example, of 3 patients with time-on-treatment 0-6 months, 2/3 were TROP2-high (IHC; e.g. case 300834, **Fig. 5D**) and 1/3 were TROP2-medium/high (inferred from RNA-seq), while of 2 patients with time-on-treatment >12 months, 1/2 had concurrent TROP2-high and TROP2-medium samples (IHC) and 1/2 was TROP2 medium/low (inferred from RNA-seq) (e.g. case 300864, **Fig. 5E**).

## Discussion

With the approval of new targeted therapies for metastatic TNBC that have overlapping indications, determining the prevalence and temporal dynamics of biomarker expression from the early stage to metastatic setting, the co-expression patterns of these markers, and their association with efficacy has become increasingly critical. Here, we report the results from comprehensive multi-platform and multi-omics profiling of a longitudinal cohort comprising more than 350 samples from 110 patients diagnosed with TNBC. We observed statistically significant decreases in both HER2 expression as determined by IHC (i.e., decrease in HER2-low, increase in HER2-0 tumors) and in TILs abundance (i.e., decrease in sTILs high/medium, increase in sTILs low) in metastatic compared to primary (DX/RD) tumor samples, with a similar trend observed for PD-L1 (i.e., decrease in PD-L1-high, increase in PD-L1-low). In contrast to HER2, TROP2 IHC expression remained relatively constant over time with approximately 90% of tumors at any given time point exhibiting TROP2 high/medium expression (H-score>100), consistent with previously reported expression in TNBC^21^. Similar distribution of TROP2 was also observed between HER2-0 vs HER2-low and between PD-L1-high vs PD-L1-low TNBC.

Instability of HER2-low status has been described among patients with HER2-negative metastatic breast cancer, with both increase and decrease in HER2 expression observed from primary to metastatic tumors, and from pre- to post-neoadjuvant tumors^18,19,27^. Multiple factors, including pre-/analytical factors, HER2 heterogeneity, and tumor evolution under therapeutic pressure, may contribute to these temporal differences in HER2 expression between sample time points. Even across different anatomic sites at the same time point, or within the same organ, synchronous HER2-low, ultra-low, and null lesions can co-exist^28^, reflecting underlying intrapatient spatial HER2 heterogeneity. In this TNBC cohort, central and local HER2 IHC assessments demonstrated approximately 80% concordance, though most discordant cases were classified as HER2-0 by local testing and HER2-low by central review, thereby identifying potential candidates for HER2-directed ADC. Given the well-documented interobserver variability with visual computer-unaided scoring^25^, and low inter-observer pathologist concordance for distinguishing HER2-0 vs HER2-low tumors^20^, improved ADC target quantification methods are urgently needed to discern expressing vs non-expressing tumors, particularly at the lower end of the spectrum of expression.

Given these inherent challenges with conventional IHC analysis, we systematically evaluated patterns of mRNA and protein expression using alternative methods, including transcriptome and exome sequencing, proteomic profiling via MS, and computational pathology quantification of protein membrane intensity. While *ERBB2* mRNA expression was significantly higher in HER2-low vs HER2-0 samples, gene expression did not clearly separate these groups. In contrast, *TACSTD2* mRNA expression was not only higher in TROP2-high/medium vs TROP2-low samples but demonstrated limited overlap in the distribution of *TACSTD2* expression between IHC groups, although the number of TROP2-low samples was limited (N=7). The strong correlation between TROP2 H-scores and *TACSTD2* expression, and limited overlap of *TACSTD2* expression between IHC groups, suggests that transcriptomic methods could serve as a reliable alternative approach to monitor and quantify levels of TROP2 expression. Since *ERBB2* expression from targeted panels has been able to separate HER2-low and HER2-0 samples^29^, the inability to clearly separate these groups in our analysis could be related to the lower resolution of transcriptome-wide RNA-seq. For both HER2 and TROP2, protein membrane staining intensity from computational pathology and protein expression measured by MS strongly correlated with IHC, distinguishing IHC groups with either limited overlap (TROP2) or reduced overlap when compared to RNA-seq data (HER2). TROP2 and HER2 quantification by MS has been used to calibrate emerging methodologies, such as quantitative immunofluorescence (QIF) assays, to characterize expression of both biomarkers^30^, and may facilitate better automated differentiation of low levels of expression compared to standard manual IHC scoring. In non-small cell lung cancer, TROP2 normalized membrane ratio (NMR; membrane SI / membrane SI + cytoplasm SI) by QCS computational pathology identified patients with longer median PFS and higher response rate with Dato-DXd.^31,32^

Given the limited number of patients who received treatment with an ADC in this cohort, it was not possible to explore correlations of ADC target expression measured across different assays with clinical outcomes with TROP2 or HER2 ADCs. In an exploratory analysis from the ASCENT trial (n=290), patients with metastatic TNBC had longer median PFS and overall survival (OS) and higher overall response rate (ORR) with SG than treatment of physician’s choice (TPC) irrespective of TROP2 IHC category, though at higher TROP2 expression there was greater magnitude of benefit^14^. For patients with TROP2 H-score <100, median PFS was 2.7 with SG vs 1.6 months with TPC, median OS, 9.3 vs 7.6 months, and ORR 22% vs 6%; for H-score 100-200: 5.6 vs 2.2 months, 14.9 vs 6.9 months, and 38% vs 11%; for H-score >200-300: 6.9 vs 2.5 months, 14.2 vs 6.9 months, and 44% vs 1%, respectively. Similarly, benefit from SG compared to TPC was observed irrespective of TROP2 protein or gene expression in the TROPiCS-02 trial. Here, among 462 patients with TROP2 IHC data, median PFS and OS in the H-score <100 group were 5.3 and 14.6 months, respectively, and in the H-score ≥100 group, 6.4 and 14.4 months. By *TACSTD2* mRNA (n=197), median PFS and OS in the group below the median (<10.5 transcripts per million) were 5.6 and 14.2 months, and in the group above the median, 7.3 and 14.4 months, with comparable ORR between TROP2 groups (19% and 21%, respectively^33,34^). It remains unclear if methods beyond IHC, immunofluorescence, or RNA-sequencing may help identify the minimum amount of target expression necessary for clinically meaningful anti-tumor activity with the newer generation of ADCs with selectively cleavable linkers. Computer-assisted quantification methods (e.g., computational assessment of NMR by QCS^24,32^) and spatial characterization of target distribution within the tumor microenvironment may provide additional information to improve patient stratification and identify those most likely to benefit from ADC therapy.

In addition to TROP2 and HER2, we evaluated immune markers of interest, PD-L1 and sTILs, in this cohort. Here, PD-L1 IHC was analyzed using the Ventana SP263 assay, with a similar PD-L1-positivity (defined as TAP ≥5%) rate of 38% in metastatic samples compared to that reported in the KN355 trial, which utilized the pharmDx 22C3 assay (PD-L1-positive defined as CPS ≥10^4^) and demonstrated a significant improvement in PFS and OS with the addition of pembrolizumab to chemotherapy in patients with previously untreated PD-L1-positive metastatic TNBC. While the distribution of *CD274* expression overlapped between PD-L1-high vs -low groups, PD-L1 protein expression by MS separated more clearly between PD-L1 IHC groups. Overall, this suggests that only markers with either high levels of expression (e.g. *ERBB2* in HER2-positive tumors) or expressed preferentially in tumor cells (e.g. *TACSTD2* in TNBC) can be accurately quantified using bulk RNA-seq, while for others, such as *ERBB2* or *CD274* in TNBC, only limited quantification resolution can be achieved. Irrespective of levels of protein expression, MS clearly was strongly correlated with IHC across markers of interest; whether target quantification by MS could improve patient stratification for ADC benefit, particularly among low-expressing tumors, remains unclear.

Consistent with other studies, sTILs abundance was significantly lower in metastatic than primary tumors^35,36^, with a similar trend observed for PD-L1 expression. In fact, among the markers evaluated in this study, only sTILs and PD-L1 were significantly correlated. Overall, 94% of PD-L1-high tumors had sTILs ≥10% (vs 64% of PD-L1-low tumors), and the higher lymphocytic infiltration among PD-L1-high tumors was observed across DX, RD, and MR time points. No statistically significant differences in HER2 or TROP2 status were identified between PD-L1 IHC groups, although numerically more tumors were HER2-low if PD-L1-low (43%) than PD-L1-high (30%). Given that samples were collected prior to the approval of pembrolizumab for high-risk early-stage TNBC (July 2021), the impact of neo-/adjuvant ICI on the dynamics of sTILs and PD-L1 across time points is unknown.

When evaluating associations with clinicopathologic features, sTILs were significantly correlated with age, due to a decrease in sTILs with age (<40 years, 83%; 40-60 years, 79%; >60 years, 57%), although this was not significant when corrected for multiple comparisons. Interestingly, this finding was driven by post-treatment samples (RD or MR) and not observed in primary DX tumors, suggesting differences by age in immune cell recruitment to the tumor microenvironment in response to cytotoxic therapy and, potentially, the ability to launch an antitumor immune response. Consistent with the previously noted association between sTILs and PD-L1, there was also a numerical decrease in PD-L1-high tumors with age (<40 years, 62%; 40-60 years, 46%; >60 years, 35%). The decrease in PD-L1 observed with age aligns with prior work, which found that, although PD-L1 tends to increase with age in most cancers, this was not the case in breast cancer, in which a small numerical decrease in PD-L1 positivity (> 5%) by IHC with age was seen in patients >60 years compared to those with <60 years^37^.

This study represents a large and comprehensive multi-omics and multi-platform profiling effort conducted across matched longitudinal (DX, RD, MR) samples in TNBC. Limitations of our study include (1) the inability to directly correlate ADC target expression with therapeutic efficacy, given that samples in this cohort were derived from patients diagnosed with metastatic TNBC prior to the first FDA approval of an ADC for TNBC (sacituzumab govitecan in April 2020); (2) that the HER2-0 and HER2-low classification is based primarily on HER IHC and not HER2 ISH, since ISH was not done for most IHC 0 and 1+ samples, and was either not done or equivocal for many IHC 2+ samples; (3) that local HER2 IHC assessment was done prior to the introduction of HER2-low as a classification with treatment implications, which could have impacted the robustness of the IHC 0 and 1+ classifications; and (4) the inability of bulk-level multi-omics analyses to distinguish tumor cell-specific versus microenvironment contributions to overall biomarker expression profiles.

With multiple ADCs and ICIs now approved for the treatment of metastatic TNBC, and additional targeted therapies in advanced clinical development, studies to help inform optimal treatment selection and combinations are critical. While ADC target expression levels to date have not identified patients with TNBC who are more likely to benefit from these agents, further investigation is needed to determine if improved methods of quantification, such as single-cell-resolution computational pathology approaches, may help discern the minimum amount of target required for antitumor activity and aid in the selection of the preferred initial ADC-based strategy in an individualized manner. Clinical trials are ongoing to assess the efficacy and safety of sequential ADCs with planned multi-omics analyses that may help elucidate if and how ADC target expression should be utilized to guide treatment selection (NCT06533826, NCT06263543). Ultimately, understanding the molecular and cellular mechanisms that drive therapeutic response and acquired resistance will be essential to inform evidence-based treatment strategies, including optimal drug combinations, sequencing approaches, and duration of therapy across breast cancer subtypes.

## Methods

Some of the methods reported follow closely what has been described in our recent work^26,38,39^.

### Compliance with Ethical Standards

The study was conducted in accordance with the International Conference on Harmonization Good Clinical Practice Standards and the Declaration of Helsinki. Institutional review board (IRB) approval was obtained at Dana-Farber/Harvard Cancer Center (DF/HCC).

### Study Design, Patient Population and Samples

Prior to any study procedures, all patients provided written informed consent for collection of de-identified clinical data, archival tissue, and/or research blood and tissue biopsies, and for sequencing of these samples, as approved by the Dana-Farber/Harvard Cancer Center IRB (DF/HCC protocols #05-246, #93-085, and secondary research use protocol #17-644). Metastatic core biopsies were obtained from patients, and samples were immediately snap-frozen in OCT and stored in −80°C. Archival formalin-fixed, paraffin-embedded (FFPE) blocks of primary and metastatic tumor samples were also obtained. A blood sample was obtained and whole blood was stored at −80°C. DNA and RNA were extracted from tumors. Germline DNA was extracted from peripheral blood mononuclear cells from whole blood or normal tissue if whole blood was not available.

Patients were identified from two sources: 1) an institutional database including all consecutive patients who underwent surgery for stage I-III breast cancer at Dana-Farber Brigham Cancer Center between 2015-2018, and 2) a prospective research biopsy protocol for patients with metastatic breast cancer. Patients were included in the present analysis if they received NAC for eTNBC, or if they were diagnosed with any stage TNBC and developed mTNBC.

Patients were classified into 3 subcohorts based on initial treatment strategy: adjuvant (Adj; early TNBC, surgery as first intervention), NAC (early TNBC, neoadjuvant chemotherapy as first intervention), and dnMBC. Four patients had asynchronous primary tumors and were classified in more than one subcohort class (Adj/NAC, Adj/dnMBC); these patients were excluded from analyses restricted to subcohort of interest.

Samples were collected from the following time points: DX, defined as the diagnostic breast biopsy (or surgery, if there was no intervening NAC); RD post-NAC (if applicable); MR, defined as the first sample collected after unresectable/distant recurrence (after eTNBC) or >30 days after initiation of the first treatment regimen for *de novo* mTNBC. For matched comparisons, if an assay was performed in more than one DX or RD sample, the earliest breast sample was considered; if there was more than one MR sample, the first MR biopsy was considered.

Of the total 110 patients, all patients had at least one TNBC biopsy, defined as estrogen receptor (ER)<10%, progesterone receptor (PR)<10%, and HER2-negative (IHC 0, IHC 1+, IHC 2+/ISH non-amplified, or IHC-unknown/ISH non-amplified). 106/110 patients had a primary tumor sample with TNBC, with 92/106 patients being ER<1%, PR<1%, and HER2-negative. For 3/110 of the remaining patients, the receptor status of all their biopsies was either TNBC (in at least one biopsy) or indeterminate (none were known to have ER≥10% or PR≥10% or HER2-positive status). For the 1/110 remaining patient, with the exception of one biopsy (ER<1%, PR 30%, HER2 IHC 1+/ISH-unknown), all biopsies were TNBC or indeterminate (none were known to have ER≥10% or PR≥10% or HER2-positive status).

### Pathologist-assessed biomarkers (PD-L1, TROP2, central HER2, local HER2, sTILs, iTILs)

#### HER2 immunohistochemistry

For local HER2 IHC, clinical pathology records were reviewed for HER2 IHC results of samples. For central HER2 IHC, available stained slides were blindly reviewed by a trained pathologist. HER2 IHC was classified as HER2-0 if IHC 0 (including HER2-null and HER2-ultra-low) and HER2-low if IHC 1+ or 2+, regardless of ISH status (with only one sample being ISH-amplified). For all samples with HER2 IHC 0 and IHC 1+, and for all but one sample with IHC 2+, ISH was non-amplified, equivocal, or not done/incomplete. For the 43 samples (from 31 patients) with HER2 IHC 2+, ISH was non-amplified in 27/43 samples, amplified in 1/43 samples, equivocal in 5/43, and missing/incomplete in 10/43 samples. All 16 samples / 14 patients with a IHC 2+/ISH amplified, equivocal, or missing/incomplete samples had a primary tumor biopsy that was HER2-negative (IHC 0, IHC 1+, or IHC 2+/ISH non-amplified).

#### TROP2 immunohistochemistry

Tumor tissue FFPE slides were stained for TROP2 using the Abcam clone EPR20043 on a Ventana Discovery platform. Tumor cell membrane TROP2 expression was scored by a trained pathologist (intensity: 0 or absent, 1+ or weak; 2+ or moderate; 3+ or strong). H-scores were calculated by adding the percentage of positive cells at 1+, 2+ (multiplied by 2), and 3+ (multiplied by 3). Samples were classified as: low (H-score <100); medium (100-200); high (>200-300), following the TROP2 analysis described in the phase III ASCENT trial^14^.

#### PD-L1 immunohistochemistry

PD-L1 expression was evaluated by staining tumor tissue FFPE slides with the ready-to-use Ventana PD-L1 (SP263) IVD on a Ventana Benchmark Ultra platform. PD-L1 staining was expressed as percentage values using three different scores: 1) TC, proportion of tumor cells expressing membrane PD-L1, equivalent to TPS (tumor proportional score); 2) IC, proportion of immune cells expressing PD-L1, membrane or cytoplasmic, relative to the tumor area (TA), similar to the SP142 Roche Scoring Method; 3) TAP (tumor area positivity), proportion of whole tumor area occupied by PD-L1 staining tumor and immune cells combined. All values are expressed as percentages with a range of 0-100; some cases are scored as <1, to indicate PD-L1 staining in some cells accounting for less than 1%. Samples were considered non-evaluable (NE) if the slide section contained less than 100 tumor cells. Samples were classified as: low (TAP <5%); high (TAP ≥5%) based on the BEGONIA 1st-line metastatic TNBC phase 1b/2 study^40^.

#### Tumor-infiltrating lymphocytes

H&E stained slides were scored for sTILs and iTILs by a trained pathologist following the International TILs Working Group guidelines^23^. Thresholds for categorization of sTILs and iTILs were based on the quartiles of their distribution, so that approximately 25% of samples were in the “low” category, 50% in the “medium” category, and 25% in the “high” category. For iTILs, given the low dynamic range, only “high” (approximately 25% of samples) and “low” categories (the remaining 75% of samples) were used. Samples were classified as: sTILs low (<10%); sTILs medium (≥10% and <40%); sTILs high (≥40%); iTILs low (<5%); iTILs high (≥5%).

### Whole exome sequence data processing and analysis

Data processing and quality control. Whole exome sequences for each tissue/blood sample were captured using Illumina technology and the sequencing data processing and analysis was performed using the Picard and Terra pipelines at the Broad Institute. The Picard pipeline (http://picard.sourceforge.net) was used to produce a BAM file with aligned reads. This includes alignment to the GRCh37 human reference sequence using the BWA aligner^41^ and estimation and recalibration of base quality score with the Genome Analysis Toolkit (GATK)^42^.

A custom-made cancer genomics analysis pipeline was used to identify somatic alterations using the Terra platform (https://app.terra.bio/). The CGA WES Characterization pipeline developed at the Broad Institute was used to call, filter and annotate somatic mutations and copy number variation. See **Supplemental Methods** for details of the variant calling pipeline used in this study. GATK CNV^43^ was used for the generation of accurate relative copy-number profiles from the whole exome sequencing data and reference/alternate read counts at heterozygous SNP sites present in both the normal and tumor samples. After accurate proportional coverage profiles are generated for a sample, Allelic CapSeg tool^44^ was used to generate a segmented allelic copy ratio profile. Allelic copy number profile and mutational call data were modeled jointly by ABSOLUTE^45^ to produce purity for the samples, a discrete copy number profile, and compute cancer cell fractions (CCF). Tumor samples with a purity of 10% or more were used for downstream analysis.

Annotating oncogenic mutations. OncoKB^46^ was used to annotate known oncogenic mutations, identify their effect (e.g. loss or gain of function) and if they are known cancer hotspots.

Corrected quantification of copy number, gene deletions, and biallelic inactivations. The inference of gene amplifications, gene deletions, and biallelic inactivations were based on the copy number profile obtained from ABSOLUTE^45^. To infer biallelic inactivations, mutational events that included both loss of heterozygosity (LOH) and a loss-of-function mutation (LOF) (loss-of-function or likely loss-of-function OncoKB-annotated mutation, or a Nonsense Mutation, Nonstop_Mutation, Frame_Shift_Del, or Frame_Shift_Del mutation) were used. Gene amplifications and deep deletions were based on the purity corrected measure for the segment containing that gene. Genes in a segment-specific copy number of less than 0.5 were considered deep deletions (Deep DEL). To measure segment-specific copy number amplifications, the genome ploidy was subtracted for each sample to obtain the copy number above ploidy (CNAP). CNAPs of at least 3 were considered as amplifications (AMP); CNAPs above 1.5 and below 3 were considered low amplification (GAIN); CNAPs of at least 6 were considered high amplifications (High AMP), and CNAPs of at least 9 and no more than 100 genes were considered focal high amplification (Focal High AMP).

### Statistical tests and analysis

Statistical significance of the enrichment between groups of variables was measured using a two-sided Fisher exact test for categorical values (biomarker status vs all clinical and sample characteristics except age group) and a two-sided Wilcoxon rank sum test for ordinal variables (biomarker status vs age group). For difference in gene expression between biomarker groups, a two-sided Welch’s t-test was used. For correlation between biomarker quantification and gene expression, Pearson correlation was used. For enrichment of longitudinal change in biomarkers of paired samples from the same patient, a two-sided paired Wilcoxon signed rank test was used between the categorical variables; % increase or decrease was calculated with respect to the categorical variables specified. For enrichment of longitudinal change in PAM50 of paired samples from the same patient, a McNemar’s Chi-squared test was used between the categorical variables. A *P* value of < 0.05 was considered statistically significant. For comparisons across all pairs of biomarkers and across multiple clinical/sample characteristics, a false discovery rate (FDR) was calculated and an FDR < 0.1 was considered statistically significant. Overlap of IQR between groups was measured as the size of the intersection of the IQRs divided by the size of smallest IQR. All statistical analysis was performed using R (version 4.3.2).

### Transcriptome sequencing data processing and analysis

#### Data processing and quality control

RNA-seq reads were mapped to the human genome (hg19) with STAR aligner^47^ with default parameters. Transcriptome quality was assessed using RNA-SeQC 2^48^ and gene expression quantification was conducted using RSEM^49^. Samples with <8,000 unique genes were removed from subsequent analysis. Gene expression was measured using upper quartile normalized log2(TPM+1) (TPM: transcripts per million) values of expressed genes and corrected for tissue type (frozen/OCT vs. FFPE) with receptor status as a covariate using ComBat^50^ in the *sva* package. Corrected and normalized gene expression with ComBat was carried out in an integrated dataset of >800 tumor samples consisting of the current cohort and other metastatic breast cancer cohorts (The Metastatic Breast Cancer Project^26^ and a Dana-Farber Cancer Institute cohort of patients with hormone receptor-positive metastatic breast cancer^38,51,52^) sequenced at Broad Institute that spans all receptor subtypes.

#### Transcriptional signature activity

To calculate the activity of a transcriptional signature or gene set, we used single-sample gene set enrichment analysis (ssGSEA). ssGSEA was performed for all tumor samples using fgsea^53^ to calculate normalized enrichment scores for Hallmark gene sets from the Molecular Signatures Database^54^ using normalized and corrected gene expression values.

#### PAM50 molecular subtype assignment

To assign research-based PAM50 subtypes, expression values were rescaled relative to those of a receptor status-balanced cohort derived from the integrated dataset, in which samples were re-sampled to achieve the ER-positive to ER-negative receptor status ratio in the UNC training set, from which the PAM50 subtype centroids were derived^55,56^. genefu was used to call research-based PAM50 subtypes^57^ using the rescaled expression values and spearman correlation to the PAM50 subtype centroids. Samples with a PAM50 centroid correlation <0.10 for each centroid were not assigned a PAM50 subtype (Not Classified, NC). Samples with PAM50 normal subtype were considered to have lower tumor purity and were removed for analyses involving RNA-seq gene expression and PAM50 subtypes.

### Image Analysis and Laser Microdissection (LMD) for proteomics by MS

A 10-µm-thick FFPE tissue section was mounted on metal-framed polyethylene naphthalate (PEN) membrane slides (Leica Microsystems) for LMD, deparaffinized with xylene, and stained with hematoxylin. A pathologist used an adjacent H&E-stained section to define the tumor regions for collection and analysis. Digital imaging of the H&E was conducted using the Leica Aperio AT2 scanner (Leica Microsystems) at 20_×_magnification. Tissue classification and image analysis was performed using Halo AI (v3.5.3577) software (Indica Labs). Tumor areas were subsequently isolated by image analysis-guided LMD using the LMD7 System (Leica Microsystems) into microcentrifuge tubes and stored in acetonitrile at -30°C until proteolytic processing.

### Proteolytic Sample Preparation for proteomics

Laser microdissected tissue samples were subjected to downstream proteolytic digestion using an automated SP3 method^58^. Tissue samples were processed in 8-strip tubes, adding lysis buffer containing 4% sodium dodecyl sulfate, 40 mM chloroacetamide, 10 mM tris(2-carboxytheyl)phosphine, and 100mM tris(hydroxymethyl)aminomethane (Tris), pH 8, before incubation at 95°C with shaking at 1,000 rpm for 1.5 hrs using an Eppendorf ThermoMixer C. Following lysis/protein de-crosslinking, lysate and tissue material were transferred into a 96-well plate and then processed using an Agilent Bravo Liquid Handling Platform (Agilent) with the following steps performed: protein conjugation onto Sera-Mag carboxylate beads (Sigma), bead washing with 80% ethanol and acetonitrile, followed by resuspension of the beads into a volume of 100mM Tris and addition of trypsin (Promega). Samples were subjected to incubation at 37°C at 1,000 rpm on a ThermoMixer overnight. Samples were acidified using trifluoroacetic acid to a final concentration of 1% using the Agilent Bravo. Recovered peptide concentration was determined using a bicinchoninic acid assay (Thermo Fisher Scientific) with 10% RIPA buffer used as a diluent.

### Parallel Reaction Monitoring Analysis LC-MS Proteomic for targeted proteomics

500 ng of peptide material were subjected to Parallel Reaction Monitoring (PRM) analysis as previously described^59^. In brief, 600 ng of peptide were combined with synthetic isotope-labelled peptides (2.4 fmol). Five-sixths of this mixture (500 ng of total peptide and 2 fmol of each synthetic peptide) were loaded onto EvoTip trapping columns before separation with the EvoSep One nanoLC system (EvoSep) coupled to an Orbitrap Exploris mass spectrometer with a FAIMS-PRO interface (Thermo Fisher). Peptides were eluted over a 44-min gradient, from 7 to 30% acetonitrile (on-column), at a flow rate of 500 nL/min. The FAIMS-PRM experiment employed higher-energy collisional dissociation (HCD) fragmentation with an isolation window of 0.7 mass-to-charge ratio (m/z), a target automatic gain control of 1E5 ions (“standard”), and a maximum injection time of 118 ms. Tandem MS (MS/MS) scans were acquired in centroid mode with the Orbitrap detector, using 60 K resolution at 200 m/z. FAIMS was operated at the standard resolution, with no additional FAIMS gas.

### Data Independent Acquisition (DIA) LC-MS proteomics

200 ng of peptide material was subjected to data independent acquisition (DIA) analysis. Samples were spiked with 1 injection equivalent of an iRT standard (Biognosys) and loaded onto EvoTip trapping column (EV2011, EvoSep) before separation with the EvoSep One nanoLC system (EvoSep) coupled to a timsTOF Pro (Bruker) mass spectrometer with a Captive spray source. Mobile phases A and B were water with 0.1% formic acid and acetonitrile with 0.1% formic acid, respectively. Peptides were separated using the 30SPD gradient and a PepSep column (C18 resin, 15 cm × 150 mm, 1.9 mm, Bruker) at a flowrate of 500 mL/min, heated to 30°C using in-line column oven (Sonation). The mass spectrometer was operated in data-independent acquisition mode using parallel accumulation-serial fragmentation (dia-PASEF)^60^. The capillary voltage was set to 1500V, with ramp and accumulation time both set to 100 ms. The collision energy was ramped linearly as a function of the mobility from 20 eV at 1/K_0_ = 0.7 Vs cm^-2^ to 52 eV at 1/K_0_ = 1.6 Vs cm^-2^. A TIMS survey scan was performed in a mass range of 100-1700 m/z, followed by twenty overlapping diaPASEF 16 m/z wide windows in a precursor mass range of 400 to 100 m/z with the Bruker Compass Hystar software estimating a cycle time of 2.23 s.

### DIA and PRM Proteomic Data Processing, Quantification and Normalization

DIA files were processed in Spectronaut (version 18.6.231227.55695) using a directDIA approach and searched against the UniProtKB Swiss-Prot human protein database (version January 2024; 20,413 reviewed sequences) using default settings. Carbamidomethyl (C) was set as a fixed modification, while Acetyl (Protein N-term) and Oxidation (M) were set as variable modifications. Stringent identification settings were used, with Precursor Q-value Cutoff, Precursor PEP Cutoff, Protein Q-value Cutoff (Experiment), Protein Q-value Cutoff (Run), and Protein PEP Cutoff were all set to 0.01. Precursors were quantified on the MS2 level and Cross-run Normalization was employed, selecting Global Normalization and Median settings.

PRM files were processed using an in-house pipeline for peak extraction and quantification from heavy:light ratio (manuscript in preparation). Fragment ions with interference were automatically flagged and reviewed by manual analysis by comparing coelution and fragment ion ratios between endogenous and reference peptides. Any fragment ions showing interference were omitted from use in quantitation. PRM protein expression was measured in amol/μg.

### Image analysis quantification methods for assessing target IHC expression and sTILs

Digitized whole-slide images of TROP2 and HER2 IHC-stained tissue were analyzed with the QCS computational pathology approach^24^: first, supervised, deep-learning model detecting the epithelial regions is followed by a second model able to perform cell detection and segment the subcellular compartments. The intensity of the staining is then quantified and provided as readouts for the related compartments of the cell. Cell-level quantifications are then aggregated at a slide level by using quantiles such as the median.

TILs were quantified using a computational pathology pipeline comprising two convolutional neural network (CNN) models for tissue segmentation and cell detection, respectively. First, a deep learning–based tissue segmentation model was applied to whole-slide images to delineate histologic compartments, with particular emphasis on separating tumor epithelium from surrounding stromal regions. The model was trained on expert-annotated data to accurately distinguish epithelial tumor from adjacent stroma, enabling precise spatial localization of the tumor microenvironment. Subsequently, a second CNN-based cell detection model was employed to reliably identify and localize individual lymphocytes within the stromal regions. Detected cells were classified based on morphological features consistent with lymphocytes and mapped to their respective tissue compartment. Quantitative readouts included total stromal area (mm²) and the number of lymphocytes detected within stromal regions. Stromal TIL density was then calculated as the number of lymphocytes per unit stromal area (cells/mm²) and used in the downstream analyses.

## Data Availability

Data that supports the findings of this study are available from the corresponding author upon reasonable request. Data requests will be reviewed by the corresponding author to determine whether the request is subject to any intellectual property or confidentiality obligations. Data sharing may require a data/material transfer agreement.

## Presentations

San Antonio Breast Cancer Symposium (December 2022; San Antonio, Texas, USA), European Society of Medical Oncology Congress (October 2023; Madrid, Spain).

## Code availability

All software and pipelines for genomic data generation are described in detail in the Methods and the Supplementary Methods. The scripts used to generate all figures and tables will be available in the associated GitHub repository upon acceptance. Additional code is available upon reasonable request from the corresponding author and may require a data/material transfer agreement.

## Acknowledgements

The authors would like to thank all the patients who contributed to this research. The authors acknowledge Kaitlyn Bifolck, a full-time employee of Dana-Farber Cancer Institute, for editorial and submission assistance.

