## Supplement for "Longitudinal multi-platform profiling reveals temporal dynamics of HER2, TROP2, PD-L1 and tumor-infiltrating lymphocytes in triple-negative breast cancer"

**Supplemental Methods**

**Supplemental References**

**Supplemental Tables (10)**

**Supplemental Figures (3)**

### Supplemental Methods

#### *Somatic alterations assessment*

A custom-made cancer genomics analysis pipeline was used to identify somatic alterations using the Terra platform (https://app.terra.bio/). We have utilized the CGA WES Characterization pipeline developed at the Broad Institute to call, filter and annotate somatic mutations and copy number variation (available in the Terra platform public workspace *broad-fc-getzlab-workflows/CGA_WES_Characterization_OpenAccess*). The pipeline employs the following tools: MuTect[1], ContEst[2], Strelka[3], Orientation Bias Filter[4], DeTiN[5], AllelicCapSeg[6], MAFPoNFilter[7], BLAT realignment filter, ABSOLUTE[8], GATK[9], GATK CNV[10], Picard Tools *CrosscheckFingerprints* and *CollectMultipleMetrics* (https://broadinstitute.github.io/picard/), Variant Effect Predictor[11], and Oncotator[12]. To annotate known oncogenic mutations, the OncoKB[13] annotator was used (https://github.com/oncokb/oncokb-annotator).

#### *DNA and RNA extraction for tumors and whole blood*

DNA extraction was performed as described previously[14]. For whole blood, DNA was extracted using magnetic bead-based chemistry in conjunction with the Chemagic MSM I instrument (Perkin Elmer) or the QIAsymphony SP instrument (Qiagen). Following red blood cell lysis, magnetic beads bound to the DNA were removed from solution using electromagnetized rods. Several wash steps followed to eliminate cell debris and protein residue from DNA bound to the magnetic beads. DNA was then eluted in TE buffer. For frozen tumor tissue, DNA and RNA were extracted simultaneously from a single frozen tissue or cell pellet sample using the AllPrep DNA/RNA Kit or DNA/RNA/miRNA Universal Kit (Qiagen). For FFPE tumor tissues, DNA and RNA were extracted simultaneously using Qiagen's AllPrep DNA/RNA FFPE Kit. All DNA was quantified using Picogreen. RNA was quantified via RiboGreen and RNA quality was measured by RNA Quality Score (RQS) value as determined by Caliper GX.

#### *Library construction (exomes)*

**Tumors and whole blood (Illumina exome bait).** DNA libraries for massively parallel sequencing were generated as described previously[14] with the following modifications: the initial genomic DNA input into the shearing step was reduced from 3 µg to 10-100 ng in 50 µL of solution. For adapter ligation, Illumina paired-end adapters were replaced with palindromic forked adapters (purchased from Integrated DNA Technologies) with unique dual indexed 8 base index molecular barcode sequences included in the adapter sequence to facilitate downstream pooling. Kapa HyperPrep reagents in 96-reaction kit format were used for end repair/A-tailing, adapter ligation, and library enrichment PCR. In addition, during the post-enrichment solid-phase reversible immobilization (SPRI) bead cleanup, elution volume was reduced to 30 µL to maximize library concentration, and a vortexing step was added to maximize the amount of template eluted.

**Tumors and whole blood (TWIST exome bait).** An aliquot of genomic DNA (100-250ng in 50µL) was used as the input into DNA fragmentation, also known as shearing. Shearing is performed acoustically using a Covaris focused-ultrasonicator, targeting 150bp fragments. Library preparation is performed using a commercially available kit provided by KAPA Biosystems (KAPA HyperPrep Kit with Library Amplification product KK8504) and IDT's duplex UMI adapters. Unique 8-base dual index sequences embedded within the p5 and p7 primers (purchased from IDT) are added during PCR. Enzymatic clean-ups are performed using Beckman Coultier AMPure XP beads with elution volumes reduced to 30µL to maximize library concentration.

#### *Solution-phase hybrid selection (exomes)*

**Tumors and whole blood (Illumina exome bait).** After library construction, hybridization and capture were performed using the relevant components of Illumina's Nextera Rapid Capture Exome Kit or TruSeq Rapid Exome Kit following the manufacturer's suggested protocol, with the following exceptions: first, all libraries within a library construction plate were pooled prior to hybridization. Second, the Midi plate from Illumina's Exome Kit was replaced with a skirted PCR plate to facilitate automation. All hybridization and capture steps were automated on the Agilent Bravo liquid handling system.

**Tumors and whole blood (TWIST exome bait).** After library construction, hybridization and capture were performed using the relevant components of IDT's XGen hybridization and wash kit and following the manufacturer's suggested protocol, with several exceptions. A set of 12-plex pre-hybridization pools were created. These pre-hybridization pools are created by equivolume pooling of the normalized libraries, Human Cot-1 and IDT XGen blocking oligos. The pre-hybridization pools undergo lyophilization using the Biotage SPE-DRY. Post lyophilization, custom exome bait (TWIST Biosciences) along with hybridization mastermix is added to the lyophilized pool prior to resuspension. Samples are incubated overnight. Library normalization and hybridization setup are performed on a Hamilton Starlet liquid handling platform, while target capture is performed on the Agilent Bravo automated platform. Post capture, a PCR is performed to amplify the capture material.

***Preparation of libraries for cluster amplification and sequencing (exomes)***

After post-capture enrichment, library pools were then quantified using quantitative PCR (KAPA Biosystems) with probes specific to the ends of the adapters; this assay was automated using Agilent's Bravo liquid handling platform. Based on qPCR quantification, libraries were normalized using a Hamilton Starlet to the required loading concentration.

***Cluster amplification and sequencing (exomes)***

**Tumors and whole blood (Illumina exome bait).** Cluster amplification of denatured templates was performed according to the manufacturer's protocol (Illumina) using HiSeq 2500 Rapid Run v1/v2, HiSeq 2500 High Output v4, HiSeq 4000 v1, or exclusion amplification cluster chemistry, and HiSeq 2500 (Rapid or High Output), HiSeq 4000, or HiSeq X flowcells. Flowcells were sequenced on HiSeq 2500 using v1 (Rapid Run flowcells) or v4 (High Output flowcells) Sequencing-by-Synthesis chemistry, v1 Sequencing-by-Synthesis chemistry for HiSeq 4000 flowcells, or v2.5 Sequencing-by-Synthesis chemistry for HiSeq X flowcells. The flowcells were then analyzed using RTA v.1.18.64 or later. Each pool of whole exome libraries was run on paired 76 bp runs, reading the dual-indexed sequences to identify molecular indices and sequenced across the number of lanes needed to meet coverage for all libraries in the pool.

**Tumors and whole blood (TWIST exome bait).** Cluster amplification of library pools was performed according to the manufacturer's protocol (Illumina) using Exclusion Amplification cluster chemistry and NovaSeq S4 flowcells. Flowcells were sequenced on Sequencing-by-Synthesis chemistry for NovaSeq S4 flowcells using paired 151bp runs.

***cDNA library construction (transcriptomes)***

**Transcriptome capture (FFPE tissue).** Total RNA was assessed for quality using the Caliper LabChip GX2. The percentage of fragments with a size greater than 200nt (DV200) was calculated using software. An aliquot of 200ng of RNA was used as the input for first strand cDNA synthesis using Illumina's TruSeq RNA Access Library Prep Kit. Synthesis of the second strand of cDNA was followed by indexed adapter ligation. Subsequent PCR amplification enriched for adapted fragments. The amplified libraries were quantified using an automated PicoGreen assay.

200 ng of each cDNA library, not including controls, were combined into 4-plex pools. Capture probes that target the exome were added and hybridized for the recommended time. Following hybridization, streptavidin magnetic beads were used to capture the library-bound probes from the previous step. Two wash steps effectively remove any nonspecifically bound products. These same hybridization, capture and wash steps are repeated to assure high specificity. A second round of amplification enriches the captured libraries. After enrichment, the libraries were quantified with qPCR using the KAPA Library Quantification Kit for Illumina Sequencing Platforms and then pooled equimolarly. The entire process is in 96-well format and all pipetting is done by either Agilent Bravo or Hamilton Starlet.

**Tru-Seq strand specific large insert RNA sequencing (frozen tissue).** Total RNA was quantified using the Quant-iT™ RiboGreen® RNA Assay Kit and normalized to 5 ng/ul. Following plating, 2 µL of ERCC controls (using a 1:1000 dilution) were spiked into each sample. An aliquot of 200 or 325 ng for each sample was transferred into library preparation which uses an automated variant of the Illumina TruSeq™ Stranded mRNA Sample Preparation Kit. This method preserves strand orientation of the RNA transcript. It uses oligo dT beads to select mRNA from the total RNA sample. It is followed by heat fragmentation and cDNA synthesis from the RNA template. The resultant 400 bp cDNA then goes through dual-indexed library preparation: 'A' base addition, adapter ligation using P7 adapters, and PCR enrichment using P5 adapters. After enrichment the libraries were quantified using Quant-iT PicoGreen (1:200 dilution). After normalizing samples to 5 ng/μL, the set was pooled and quantified using the KAPA Library Quantification Kit for Illumina Sequencing Platforms. The entire process is in 96-well format and all pipetting is done by either Agilent Bravo or Hamilton Starlet.

#### *Illumina sequencing (transcriptomes)*

Pooled libraries were normalized to 2 nM and denatured using 0.1 N NaOH prior to sequencing. Flowcell cluster amplification and sequencing were performed according to the manufacturer's protocols using HiSeq 2000, HiSeq 2500, or NovaSeq 6000. Each run was a 76 (Transcriptome capture) or 101 (Tru-Seq) bp paired-end with an eight-base index barcode read. Data were analyzed using the Broad Picard Pipeline which includes de-multiplexing and data aggregation.

#### *Figure and plot generation and editing*

Plots were generated using R and the packages ggplot2[15], ggpubr[16], and ComplexHeatmap[17]. Figures were edited using Microsoft PowerPoint and Inkscape.

**Supplemental Table S1. Prevalence of biomarker status (pathologist assessment) at different time points in each subcohort (adjuvant, neoadjuvant, de *novo* MBC).** One sample per patient for each time point are included. DX, at diagnosis; RD, residual disease; MR, metastatic/recurrence; Adj, adjuvant (surgery as first intervention for early TNBC); NAC, neoadjuvant (chemotherapy as first intervention for early TNBC); dnMBC (*de novo* metastatic TNBC). L-IHC, local IHC.

| **Biomarker status / Sample timepoint** | **DX** | **RD** | **MR** | **Total** |
|---|---|---|---|---|
| **Adj HER2 L-IHC (N=37, 20 patients)** | | | | |
| 0 | 9 (40.9%) | 0 (0.0%) | 13 (59.1%) | 22 (100.0%) |
| 1+ | 5 (55.6%) | 0 (0.0%) | 4 (44.4%) | 9 (100.0%) |
| 2+ | 3 (50.0%) | 0 (0.0%) | 3 (50.0%) | 6 (100.0%) |
| 3+ | 0 (-) | 0 (-) | 0 (-) | 0 (100.0%) |
| Total | 17 (45.9%) | 0 (0.0%) | 20 (54.1%) | 37 (100.0%) |
| **Adj TROP2 status (N=23, 19 patients)** | | | | |
| Low | 2 (66.7%) | 0 (0.0%) | 1 (33.3%) | 3 (100.0%) |
| Medium | 6 (66.7%) | 0 (0.0%) | 3 (33.3%) | 9 (100.0%) |
| High | 3 (27.3%) | 0 (0.0%) | 8 (72.7%) | 11 (100.0%) |
| Total | 11 (47.8%) | 0 (0.0%) | 12 (52.2%) | 23 (100.0%) |
| **Adj PD-L1 status (N=19, 14 patients)** | | | | |
| Low | 5 (50.0%) | 0 (0.0%) | 5 (50.0%) | 10 (100.0%) |
| High | 5 (55.6%) | 0 (0.0%) | 4 (44.4%) | 9 (100.0%) |
| Total | 10 (52.6%) | 0 (0.0%) | 9 (47.4%) | 19 (100.0%) |
| **Adj sTILs status (N=26, 18 patients)** | | | | |
| Low | 3 (33.3%) | 0 (0.0%) | 6 (66.7%) | 9 (100.0%) |
| Medium | 5 (45.5%) | 0 (0.0%) | 6 (54.5%) | 11 (100.0%) |
| High | 4 (66.7%) | 0 (0.0%) | 2 (33.3%) | 6 (100.0%) |
| Total | 12 (46.2%) | 0 (0.0%) | 14 (53.8%) | 26 (100.0%) |
| **NAC HER2 L-IHC (N=149, 76 patients)** | | | | |
| 0 | 33 (37.5%) | 31 (35.2%) | 24 (27.3%) | 88 (100.0%) |
| 1+ | 26 (66.7%) | 10 (25.6%) | 3 (7.7%) | 39 (100.0%) |
| 2+ | 10 (47.6%) | 9 (42.9%) | 2 (9.5%) | 21 (100.0%) |
| 3+ | 0 (0.0%) | 1 (100.0%) | 0 (0.0%) | 1 (100.0%) |
| Total | 69 (46.3%) | 51 (34.2%) | 29 (19.5%) | 149 (100.0%) |
| **NAC TROP2 status (N=92, 62 patients)** | | | | |
| Low | 3 (33.3%) | 4 (44.4%) | 2 (22.2%) | 9 (100.0%) |
| Medium | 8 (28.6%) | 16 (57.1%) | 4 (14.3%) | 28 (100.0%) |
| High | 23 (41.8%) | 24 (43.6%) | 8 (14.5%) | 55 (100.0%) |
| Total | 34 (37.0%) | 44 (47.8%) | 14 (15.2%) | 92 (100.0%) |
| **NAC PD-L1 status (N=74, 57 patients)** | | | | |
| Low | 11 (30.6%) | 19 (52.8%) | 6 (16.7%) | 36 (100.0%) |
| High | 14 (36.8%) | 20 (52.6%) | 4 (10.5%) | 38 (100.0%) |
| Total | 25 (33.8%) | 39 (52.7%) | 10 (13.5%) | 74 (100.0%) |
| **NAC sTILs status (N=110, 68 patients)** | | | | |
| Low | 4 (20.0%) | 9 (45.0%) | 7 (35.0%) | 20 (100.0%) |
| Medium | 27 (45.8%) | 28 (47.5%) | 4 (6.8%) | 59 (100.0%) |
| High | 16 (51.6%) | 11 (35.5%) | 4 (12.9%) | 31 (100.0%) |
| Total | 47 (42.7%) | 48 (43.6%) | 15 (13.6%) | 110 (100.0%) |
| **dnMBC HER2 L-IHC (N=16, 10 patients)** | | | | |
| 0 | 5 (45.5%) | 0 (0.0%) | 6 (54.5%) | 11 (100.0%) |
| 1+ | 2 (66.7%) | 0 (0.0%) | 1 (33.3%) | 3 (100.0%) |
| 2+ | 1 (50.0%) | 0 (0.0%) | 1 (50.0%) | 2 (100.0%) |
| 3+ | 0 (-) | 0 (-) | 0 (-) | 0 (100.0%) |
| Total | 8 (50.0%) | 0 (0.0%) | 8 (50.0%) | 16 (100.0%) |
| **dnMBC TROP2 status (N=9, 8 patients)** | | | | |
| Low | 0 (0.0%) | 0 (0.0%) | 1 (100.0%) | 1 (100.0%) |
| Medium | 3 (100.0%) | 0 (0.0%) | 0 (0.0%) | 3 (100.0%) |
| High | 2 (40.0%) | 0 (0.0%) | 3 (60.0%) | 5 (100.0%) |
| Total | 5 (55.6%) | 0 (0.0%) | 4 (44.4%) | 9 (100.0%) |
| **dnMBC PD-L1 status (N=7, 6 patients)** | | | | |

| | | | | |
|---|---|---|---|---|
| Low | 2 (50.0%) | 0 (0.0%) | 2 (50.0%) | 4 (100.0%) |
| High | 2 (66.7%) | 0 (0.0%) | 1 (33.3%) | 3 (100.0%) |
| Total | 4 (57.1%) | 0 (0.0%) | 3 (42.9%) | 7 (100.0%) |
| **dnMBC sTILs status (N=9, 7 patients)** | | | | |
| Low | 1 (25.0%) | 0 (0.0%) | 3 (75.0%) | 4 (100.0%) |
| Medium | 2 (66.7%) | 0 (0.0%) | 1 (33.3%) | 3 (100.0%) |
| High | 2 (100.0%) | 0 (0.0%) | 0 (0.0%) | 2 (100.0%) |
| Total | 5 (55.6%) | 0 (0.0%) | 4 (44.4%) | 9 (100.0%) |

**Supplemental Table S2. Longitudinal changes in biomarker status (pathologist assessment) between different time points in each subcohort (adjuvant, neoadjuvant, de novo MBC).** DX, at diagnosis; RD, residual disease; MR, metastatic/recurrence; Adj, adjuvant (surgery as first intervention for early TNBC); NAC, neoadjuvant (chemotherapy as first intervention for early TNBC); dnMBC (de novo metastatic TNBC). L-IHC, local IHC.

| **Biomarker status DX/RD** | **Biomarker status MR** | | | | |
|---|---|---|---|---|---|
| **Adj HER2 L-IHC status (N=34, 17 patients)** | 0 | 1+ | 2+ | 3+ | Total |
| 0 | 7 (77.8%) | 2 (22.2%) | 0 (0.0%) | 0 (0.0%) | 9 (100.0%) |
| 1+ | 3 (60.0%) | 0 (0.0%) | 2 (40.0%) | 0 (0.0%) | 5 (100.0%) |
| 2+ | 1 (33.3%) | 1 (33.3%) | 1 (33.3%) | 0 (0.0%) | 3 (100.0%) |
| 3+ | 0 (-) | 0 (-) | 0 (-) | 0 (-) | 0 (100.0%) |
| Total | 11 (64.7%) | 3 (17.6%) | 3 (17.6%) | 0 (0.0%) | 17 (100.0%) |
| **Adj TROP2 status (N=8, 4 patients)** | Low | Medium | High | Total | |
| Low | 0 (-) | 0 (-) | 0 (-) | 0 (100.0%) | |
| Medium | 0 (0.0%) | 0 (0.0%) | 2 (100.0%) | 2 (100.0%) | |
| High | 0 (0.0%) | 0 (0.0%) | 2 (100.0%) | 2 (100.0%) | |
| Total | 0 (0.0%) | 0 (0.0%) | 4 (100.0%) | 4 (100.0%) | |
| **Adj PD-L1 status (N=10, 5 patients)** | Low | High | Total | | |
| Low | 3 (100.0%) | 0 (0.0%) | 3 (100.0%) | | |
| High | 1 (50.0%) | 1 (50.0%) | 2 (100.0%) | | |
| Total | 4 (80.0%) | 1 (20.0%) | 5 (100.0%) | | |
| **Adj sTILs status (N=16, 8 patients)** | Low | Medium | High | Total | |
| Low | 1 (50.0%) | 1 (50.0%) | 0 (0.0%) | 2 (100.0%) | |
| Medium | 2 (50.0%) | 2 (50.0%) | 0 (0.0%) | 4 (100.0%) | |
| High | 2 (100.0%) | 0 (0.0%) | 0 (0.0%) | 2 (100.0%) | |
| Total | 5 (62.5%) | 3 (37.5%) | 0 (0.0%) | 8 (100.0%) | |
| **NAC HER2 L-IHC status (N=62, 31 patients)** | 0 | 1+ | 2+ | 3+ | Total |
| 0 | 11 (84.6%) | 2 (15.4%) | 0 (0.0%) | 0 (0.0%) | 13 (100.0%) |
| 1+ | 13 (76.5%) | 2 (11.8%) | 2 (11.8%) | 0 (0.0%) | 17 (100.0%) |
| 2+ | 1 (100.0%) | 0 (0.0%) | 0 (0.0%) | 0 (0.0%) | 1 (100.0%) |
| 3+ | 0 (-) | 0 (-) | 0 (-) | 0 (-) | 0 (100.0%) |
| Total | 25 (80.6%) | 4 (12.9%) | 2 (6.5%) | 0 (0.0%) | 31 (100.0%) |
| **NAC TROP2 status (N=26, 13 patients)** | Low | Medium | High | Total | |
| Low | 0 (0.0%) | 1 (100.0%) | 0 (0.0%) | 1 (100.0%) | |
| Medium | 0 (0.0%) | 1 (25.0%) | 3 (75.0%) | 4 (100.0%) | |
| High | 2 (25.0%) | 2 (25.0%) | 4 (50.0%) | 8 (100.0%) | |
| Total | 2 (15.4%) | 4 (30.8%) | 7 (53.8%) | 13 (100.0%) | |
| **NAC PD-L1 status (N=16, 8 patients)** | Low | High | Total | | |
| Low | 4 (100.0%) | 0 (0.0%) | 4 (100.0%) | | |
| High | 1 (25.0%) | 3 (75.0%) | 4 (100.0%) | | |
| Total | 5 (62.5%) | 3 (37.5%) | 8 (100.0%) | | |
| **NAC sTILs status (N=26, 13 patients)** | Low | Medium | High | Total | |
| Low | 2 (100.0%) | 0 (0.0%) | 0 (0.0%) | 2 (100.0%) | |
| Medium | 5 (62.5%) | 3 (37.5%) | 0 (0.0%) | 8 (100.0%) | |
| High | 0 (0.0%) | 0 (0.0%) | 3 (100.0%) | 3 (100.0%) | |
| Total | 7 (53.8%) | 3 (23.1%) | 3 (23.1%) | 13 (100.0%) | |
| **dnMBC HER2 L-IHC status (N=14, 7 patients)** | 0 | 1+ | 2+ | 3+ | Total |
| 0 | 5 (100.0%) | 0 (0.0%) | 0 (0.0%) | 0 (0.0%) | 5 (100.0%) |
| 1+ | 0 (0.0%) | 1 (100.0%) | 0 (0.0%) | 0 (0.0%) | 1 (100.0%) |
| 2+ | 0 (0.0%) | 0 (0.0%) | 1 (100.0%) | 0 (0.0%) | 1 (100.0%) |
| 3+ | 0 (-) | 0 (-) | 0 (-) | 0 (-) | 0 (100.0%) |
| Total | 5 (71.4%) | 1 (14.3%) | 1 (14.3%) | 0 (0.0%) | 7 (100.0%) |
| **dnMBC TROP2 status (N=2, 1 patients)** | Low | Medium | High | Total | |
| Low | 0 (-) | 0 (-) | 0 (-) | 0 (100.0%) | |
| Medium | 1 (100.0%) | 0 (0.0%) | 0 (0.0%) | 1 (100.0%) | |
| High | 0 (-) | 0 (-) | 0 (-) | 0 (100.0%) | |
| Total | 1 (100.0%) | 0 (0.0%) | 0 (0.0%) | 1 (100.0%) | |

| **dnMBC PD-L1 status (N=2, 1 patients)** | Low | High | Total | |
|---|---|---|---|---|
| Low | 0 (-) | 0 (-) | 0 (100.0%) | |
| High | 1 (100.0%) | 0 (0.0%) | 1 (100.0%) | |
| Total | 1 (100.0%) | 0 (0.0%) | 1 (100.0%) | |
| **dnMBC sTILs status (N=4, 2 patients)** | Low | Medium | High | Total |
| Low | 1 (100.0%) | 0 (0.0%) | 0 (0.0%) | 1 (100.0%) |
| Medium | 1 (100.0%) | 0 (0.0%) | 0 (0.0%) | 1 (100.0%) |
| High | 0 (-) | 0 (-) | 0 (-) | 0 (100.0%) |
| Total | 2 (100.0%) | 0 (0.0%) | 0 (0.0%) | 2 (100.0%) |

**Supplemental Table S3. Comparison of HER2 status between central and local IHC (pathologist assessment) in TNBC.** N=116 samples from 71 patients assesed with local IHC (from pathology records) and central IHC (from central pathologist review).

| | **HER2 local IHC** | | | | |
|---|---|---|---|---|---|
| **HER2 central IHC/HER2 local IHC** | 0 | 1+ | 2+ | 3+ | Total |
| 0 | 67 (57.8%) | 5 (4.3%) | 7 (6.0%) | 0 (0.0%) | 79 (68.1%) |
| 1+ | 12 (10.3%) | 9 (7.8%) | 5 (4.3%) | 0 (0.0%) | 26 (22.4%) |
| 2+ | 1 (0.9%) | 3 (2.6%) | 7 (6.0%) | 0 (0.0%) | 11 (9.5%) |
| 3+ | 0 (0.0%) | 0 (0.0%) | 0 (0.0%) | 0 (0.0%) | 0 (0.0%) |
| Total | 80 (69.0%) | 17 (14.7%) | 19 (16.4%) | 0 (0.0%) | 116 (100.0%) |

**Supplemental Table S4. Comparison of sTILs and iTILs status (pathologist assessment).** N=184 samples from 97 patients assesed for sTILs and iTILs. *P* values are a two-sided Fisher exact test between iTILs low vs high and sTILs low vs medium/high.

| | **iTILs status** | | | |
|---|---|---|---|---|
| **sTILs status** | Low | High | Total | *P* |
| Low | 48 (98.0%) | 1 (2.0%) | 49 (100.0%) | 7.78E-06 |
| Medium | 73 (79.3%) | 19 (20.7%) | 92 (100.0%) | |
| High | 21 (48.8%) | 22 (51.2%) | 43 (100.0%) | |
| Total | 142 (77.2%) | 42 (22.8%) | 184 (100.0%) | |

**Supplemental Table S5. Correlation across biomarker status (pathologist assessment).** *P* values are a two-sided Fisher exact test between biomarker status variables using the groups in Figure 3: HER2 status (0, low), TROP2 status (low, medium/high), PD-L1 status (low, medium), and sTILs status (low, medium/high). False discovery rates (FDR) are calculated using the *P* values of all distinct comparisons in this table. L-IHC, local IHC.

| **Biomarker status / HER2 L-IHC** | 0 | 1+ | 2+ | 3+ | Total | *P* | *FDR* |
|---|---|---|---|---|---|---|---|
| **TROP2 status (N=126, 86 patients)** | | | | | | | |
| Low | 9 (69.2%) | 2 (15.4%) | 2 (15.4%) | 0 (0.0%) | 13 (100.0%) | 0.767 | 1.000 |
| Medium | 27 (65.9%) | 9 (22.0%) | 5 (12.2%) | 0 (0.0%) | 41 (100.0%) | | |
| High | 44 (61.1%) | 14 (19.4%) | 14 (19.4%) | 0 (0.0%) | 72 (100.0%) | | |
| Total | 80 (63.5%) | 25 (19.8%) | 21 (16.7%) | 0 (0.0%) | 126 (100.0%) | | |
| **PD-L1 status (N=97, 72 patients)** | | | | | | | |
| Low | 28 (54.9%) | 12 (23.5%) | 10 (19.6%) | 1 (2.0%) | 51 (100.0%) | 0.208 | 1.000 |
| High | 32 (69.6%) | 7 (15.2%) | 7 (15.2%) | 0 (0.0%) | 46 (100.0%) | | |
| Total | 60 (61.9%) | 19 (19.6%) | 17 (17.5%) | 1 (1.0%) | 97 (100.0%) | | |
| **sTILs status (N=147, 90 patients)** | | | | | | | |
| Low | 21 (60.0%) | 6 (17.1%) | 8 (22.9%) | 0 (0.0%) | 35 (100.0%) | 0.543 | 1.000 |
| Medium | 51 (66.2%) | 18 (23.4%) | 7 (9.1%) | 1 (1.3%) | 77 (100.0%) | | |
| High | 23 (65.7%) | 5 (14.3%) | 7 (20.0%) | 0 (0.0%) | 35 (100.0%) | | |
| Total | 95 (64.6%) | 29 (19.7%) | 22 (15.0%) | 1 (0.7%) | 147 (100.0%) | | |
| **Biomarker status / TROP2 status** | Low | Medium | High | Total | | *P* | *FDR* |
| **HER2 L-IHC (N=126, 86 patients)** | | | | | | | |
| 0 | 9 (11.3%) | 27 (33.8%) | 44 (55.0%) | 80 (100.0%) | | 0.767 | 1.000 |
| 1+ | 2 (8.0%) | 9 (36.0%) | 14 (56.0%) | 25 (100.0%) | | | |
| 2+ | 2 (9.5%) | 5 (23.8%) | 14 (66.7%) | 21 (100.0%) | | | |
| 3+ | 0 (-) | 0 (-) | 0 (-) | 0 (100.0%) | | | |
| Total | 13 (10.3%) | 41 (32.5%) | 72 (57.1%) | 126 (100.0%) | | | |
| **PD-L1 status (N=113, 79 patients)** | | | | | | | |
| Low | 6 (10.2%) | 20 (33.9%) | 33 (55.9%) | 59 (100.0%) | | 0.571 | 1.000 |
| High | 8 (14.8%) | 16 (29.6%) | 30 (55.6%) | 54 (100.0%) | | | |

| | | | | | | | |
|---|---|---|---|---|---|---|---|
| Total | 14 (12.4%) | 36 (31.9%) | 63 (55.8%) | 113 (100.0%) | | | |
| **sTILs status (N=145, 89 patients)** | | | | | | | |
| Low | 6 (15.8%) | 9 (23.7%) | 23 (60.5%) | 38 (100.0%) | | 0.221 | 1.000 |
| Medium | 8 (10.8%) | 27 (36.5%) | 39 (52.7%) | 74 (100.0%) | | | |
| High | 1 (3.0%) | 12 (36.4%) | 20 (60.6%) | 33 (100.0%) | | | |
| Total | 15 (10.3%) | 48 (33.1%) | 82 (56.6%) | 145 (100.0%) | | | |
| **Biomarker status / PD-L1 status** | Low | High | Total | | | *P* | *FDR* |
| **HER2 L-IHC (N=97, 72 patients)** | | | | | | | |
| 0 | 28 (46.7%) | 32 (53.3%) | 60 (100.0%) | | | 0.208 | 1.000 |
| 1+ | 12 (63.2%) | 7 (36.8%) | 19 (100.0%) | | | | |
| 2+ | 10 (58.8%) | 7 (41.2%) | 17 (100.0%) | | | | |
| 3+ | 1 (100.0%) | 0 (0.0%) | 1 (100.0%) | | | | |
| Total | 51 (52.6%) | 46 (47.4%) | 97 (100.0%) | | | | |
| **TROP2 status (N=113, 79 patients)** | | | | | | | |
| Low | 6 (42.9%) | 8 (57.1%) | 14 (100.0%) | | | 0.571 | 1.000 |
| Medium | 20 (55.6%) | 16 (44.4%) | 36 (100.0%) | | | | |
| High | 33 (52.4%) | 30 (47.6%) | 63 (100.0%) | | | | |
| Total | 59 (52.2%) | 54 (47.8%) | 113 (100.0%) | | | | |
| **sTILs status (N=110, 77 patients)** | | | | | | | |
| Low | 20 (87.0%) | 3 (13.0%) | 23 (100.0%) | | | 1.07E-04 | 6.44E-04 |
| Medium | 33 (53.2%) | 29 (46.8%) | 62 (100.0%) | | | | |
| High | 3 (12.0%) | 22 (88.0%) | 25 (100.0%) | | | | |
| Total | 56 (50.9%) | 54 (49.1%) | 110 (100.0%) | | | | |
| **Biomarker status / sTILs status** | Low | Medium | High | Total | | *P* | *FDR* |
| **HER2 L-IHC (N=147, 90 patients)** | | | | | | | |
| 0 | 21 (22.1%) | 51 (53.7%) | 23 (24.2%) | 95 (100.0%) | | 0.543 | 1.000 |
| 1+ | 6 (20.7%) | 18 (62.1%) | 5 (17.2%) | 29 (100.0%) | | | |
| 2+ | 8 (36.4%) | 7 (31.8%) | 7 (31.8%) | 22 (100.0%) | | | |

| | | | | | | | |
|---|---|---|---|---|---|---|---|
| 3+ | 0 (0.0%) | 1 (100.0%) | 0 (0.0%) | 1 (100.0%) | | | |
| Total | 35 (23.8%) | 77 (52.4%) | 35 (23.8%) | 147 (100.0%) | | | |
| **TROP2 status (N=145, 89 patients)** | | | | | | | |
| Low | 6 (40.0%) | 8 (53.3%) | 1 (6.7%) | 15 (100.0%) | | 0.221 | 1.000 |
| Medium | 9 (18.8%) | 27 (56.3%) | 12 (25.0%) | 48 (100.0%) | | | |
| High | 23 (28.0%) | 39 (47.6%) | 20 (24.4%) | 82 (100.0%) | | | |
| Total | 38 (26.2%) | 74 (51.0%) | 33 (22.8%) | 145 (100.0%) | | | |
| **PD-L1 status (N=110, 77 patients)** | | | | | | | |
| Low | 20 (35.7%) | 33 (58.9%) | 3 (5.4%) | 56 (100.0%) | | 1.07E-04 | 6.44E-04 |
| High | 3 (5.6%) | 29 (53.7%) | 22 (40.7%) | 54 (100.0%) | | | |
| Total | 23 (20.9%) | 62 (56.4%) | 25 (22.7%) | 110 (100.0%) | | | |

**Supplemental Table S6. Correlation between biomarker status (pathologist assessment) and clinical characteristics.** One sample per patient for each time point (diagnosis, residual disease, metastatic/recurrence) are included. *P* values are either two-sided Fisher exact test (for the categorical clinical variables: race, stage, metastatic presentation, germline BRCA1/2 status, or subcohort) or two-sided Wilcoxon rank sum test (for the ordinal variable age) between the biomarker and clinical groups specified. False discovery rates (FDR) are calculated using the *P* values in the biomarker status and clinical/sample characteristics comparisons of Supplemental Table S5 and S6. *Germline BRCA1/2 WT status excludes BRCA1/2 VUS (N=3 pts). L-IHC, local IHC.

| **Biomarker status and clinical variable** | Biomarker groups | Clinical groups | N biomarker groups | N clinical groups | *P* | *FDR* |
|---|---|---|---|---|---|---|
| **HER2 L-IHC (N=210, 110 patients)** | | | | | | |
| Race | 0 vs Low | Black or African American vs White | 0 (N=113), Low (N=82) | White (N=174), Black or African American (N=21) | 0.244 | 0.705 |
| Age | 0 vs Low | <40 vs 40-60 vs >60 | 0 (N=123), Low (N=87) | <40 (N=52), 40-60 (N=127), >60 (N=31) | 0.281 | 0.715 |
| Stage | 0 vs Low | I vs II vs III vs IV | 0 (N=118), Low (N=83) | I (N=20), II (N=125), III (N=40), IV (N=16) | 0.679 | 1.000 |
| Metastatic presentation | 0 vs Low | *De novo* vs Recurrent | 0 (N=83), Low (N=53) | *De novo* (N=16), Recurrent (N=120) | 0.593 | 0.933 |
| BRCA1/2 status | 0 vs Low | BRCA1/2 pathogenic variant vs BRCA1/2 wild-type (WT)* | 0 (N=117), Low (N=81) | BRCA1/2 pathogenic variant (N=18), BRCA1/2 WT (N=180) | 0.317 | 0.739 |
| Subcohort | 0 vs Low | Adj vs NAC vs dnMBC | 0 (N=121), Low (N=80) | Adj (N=37), NAC (N=148), dnMBC (N=16) | 0.805 | 1.000 |
| **TROP2 status (N=130, 92 patients)** | | | | | | |
| Race | Low vs Medium/High | Black or African American vs White | Low (N=11), Medium/High (N=109) | Black or African American (N=12), White (N=108) | 0.600 | 0.933 |
| Age | Low vs Medium/High | <40 vs 40-60 vs >60 | Low (N=13), Medium/High (N=117) | <40 (N=28), 40-60 (N=82), >60 (N=20) | 0.721 | 1.000 |
| Stage | Low vs Medium/High | I vs II vs III vs IV | Low (N=13), Medium/High (N=111) | I (N=10), II (N=74), III (N=31), IV (N=9) | 0.080 | 0.553 |
| Metastatic presentation | Low vs Medium/High | *De novo* vs Recurrent | Low (N=10), Medium/High (N=78) | *De novo* (N=9), Recurrent (N=79) | 1.000 | 1.000 |
| BRCA1/2 status | Low vs Medium/High | BRCA1/2 pathogenic variant vs BRCA1/2 wild-type (WT)* | Low (N=13), Medium/High (N=109) | BRCA1/2 pathogenic variant (N=11), BRCA1/2 WT (N=111) | 1.000 | 1.000 |
| Subcohort | Low vs Medium/High | Adj vs NAC vs dnMBC | Low (N=13), Medium/High (N=111) | Adj (N=23), NAC (N=92), dnMBC (N=9) | 0.880 | 1.000 |
| **PDL1 status (N=105, 80 patients)** | | | | | | |
| Race | Low vs High | Black or African American vs White | Low (N=52), High (N=46) | Black or African American (N=10), White (N=88) | 0.508 | 0.890 |

| | | | | | | |
|---|---|---|---|---|---|---|
| Age | Low vs High | <40 vs 40-60 vs >60 | Low (N=55), High (N=50) | <40 (N=21), 40-60 (N=67), >60 (N=17) | 0.099 | 0.553 |
| Stage | Low vs High | I vs II vs III vs IV | Low (N=53), High (N=48) | I (N=10), II (N=63), III (N=21), IV (N=7) | 0.818 | 1.000 |
| Metastatic presentation | Low vs High | *De novo* vs Recurrent | Low (N=39), High (N=28) | *De novo* (N=7), Recurrent (N=60) | 1.000 | 1.000 |
| BRCA1/2 status | Low vs High | BRCA1/2 pathogenic variant vs BRCA1/2 wild-type (WT)* | Low (N=52), High (N=47) | BRCA1/2 pathogenic variant (N=7), BRCA1/2 WT (N=92) | 0.252 | 0.705 |
| Subcohort | Low vs High | Adj vs NAC vs dnMBC | Low (N=50), High (N=50) | Adj (N=19), NAC (N=74), dnMBC (N=7) | 0.888 | 1.000 |
| **sTILs status (N=152, 97 patients)** | | | | | | |
| Race | Low vs Medium/High | Black or African American vs White | Low (N=32), Medium/High (N=107) | Black or African American (N=13), White (N=126) | 1.000 | 1.000 |
| Age | Low vs Medium/High | <40 vs 40-60 vs >60 | Low (N=36), Medium/High (N=116) | <40 (N=35), 40-60 (N=94), >60 (N=23) | 0.038 | 0.421 |
| Stage | Low vs Medium/High | I vs II vs III vs IV | Low (N=34), Medium/High (N=111) | I (N=14), II (N=92), III (N=30), IV (N=9) | 0.459 | 0.890 |
| Metastatic presentation | Low vs Medium/High | *De novo* vs Recurrent | Low (N=28), Medium/High (N=68) | *De novo* (N=9), Recurrent (N=87) | 0.441 | 0.890 |
| BRCA1/2 status | Low vs Medium/High | BRCA1/2 pathogenic variant vs BRCA1/2 wild-type (WT)* | Low (N=35), Medium/High (N=107) | BRCA1/2 pathogenic variant (N=13), BRCA1/2 WT (N=129) | 0.186 | 0.667 |
| Subcohort | Low vs Medium/High | Adj vs NAC vs dnMBC | Low (N=33), Medium/High (N=112) | Adj (N=26), NAC (N=110), dnMBC (N=9) | 0.045 | 0.421 |

**Supplemental Table S7. Correlation between biomarker status (pathologist assessment) and research-based PAM50 status.** *P* values are a two-sided Fisher exact test between Basal and non-Basal PAM50 and the biomarker variables (HER2 0 vs low, TROP2 low vs medium/high, PD-L1 low vs high, and sTILs low vs medium/high). L-IHC, local IHC.

| **Biomarker status / PAM50 subtype** | Basal | HER2-enriched | Luminal A | Luminal B | Not Classified | Normal | Total | *P* |
|---|---|---|---|---|---|---|---|---|
| **HER2 L-IHC (N=78, 60 patients)** | | | | | | | | |
| 0 | 41 (78.8%) | 3 (5.8%) | 0 (0.0%) | 1 (1.9%) | 1 (1.9%) | 6 (11.5%) | 52 (100.0%) | 0.086 |
| 1+ | 11 (64.7%) | 3 (17.6%) | 0 (0.0%) | 0 (0.0%) | 0 (0.0%) | 3 (17.6%) | 17 (100.0%) | |
| 2+ | 4 (50.0%) | 2 (25.0%) | 1 (12.5%) | 0 (0.0%) | 0 (0.0%) | 1 (12.5%) | 8 (100.0%) | |
| 3+ | 0 (0.0%) | 0 (0.0%) | 0 (0.0%) | 0 (0.0%) | 0 (0.0%) | 1 (100.0%) | 1 (100.0%) | |
| Total | 56 (71.8%) | 8 (10.3%) | 1 (1.3%) | 1 (1.3%) | 1 (1.3%) | 11 (14.1%) | 78 (100.0%) | |
| **TROP2 status (N=62, 50 patients)** | | | | | | | | |
| Low | 6 (85.7%) | 1 (14.3%) | 0 (0.0%) | 0 (0.0%) | 0 (0.0%) | 0 (0.0%) | 7 (100.0%) | 1.000 |
| Medium | 11 (68.8%) | 1 (6.3%) | 1 (6.3%) | 0 (0.0%) | 1 (6.3%) | 2 (12.5%) | 16 (100.0%) | |
| High | 22 (56.4%) | 5 (12.8%) | 1 (2.6%) | 2 (5.1%) | 0 (0.0%) | 9 (23.1%) | 39 (100.0%) | |
| Total | 39 (62.9%) | 7 (11.3%) | 2 (3.2%) | 2 (3.2%) | 1 (1.6%) | 11 (17.7%) | 62 (100.0%) | |
| **PD-L1 status (N=49, 44 patients)** | | | | | | | | |
| Low | 19 (65.5%) | 4 (13.8%) | 1 (3.4%) | 0 (0.0%) | 1 (3.4%) | 4 (13.8%) | 29 (100.0%) | 0.215 |
| High | 15 (75.0%) | 0 (0.0%) | 0 (0.0%) | 1 (5.0%) | 0 (0.0%) | 4 (20.0%) | 20 (100.0%) | |
| Total | 34 (69.4%) | 4 (8.2%) | 1 (2.0%) | 1 (2.0%) | 1 (2.0%) | 8 (16.3%) | 49 (100.0%) | |
| **sTILs status (N=71, 57 patients)** | | | | | | | | |
| Low | 15 (68.2%) | 4 (18.2%) | 1 (4.5%) | 0 (0.0%) | 1 (4.5%) | 1 (4.5%) | 22 (100.0%) | 0.513 |
| Medium | 19 (57.6%) | 5 (15.2%) | 0 (0.0%) | 1 (3.0%) | 0 (0.0%) | 8 (24.2%) | 33 (100.0%) | |
| High | 12 (75.0%) | 1 (6.3%) | 0 (0.0%) | 0 (0.0%) | 0 (0.0%) | 3 (18.8%) | 16 (100.0%) | |
| Total | 46 (64.8%) | 10 (14.1%) | 1 (1.4%) | 1 (1.4%) | 1 (1.4%) | 12 (16.9%) | 71 (100.0%) | |

**Supplemental Table S8. Correlation between biomarker status (pathologist assessment) and tumor site.** N=345 samples from 110 patients with biomarker data for at least one sample: breast (N=211), axillary lymph node (N=55), chest wall (N=24), liver (N=19), other (N=45). *P* values are two-sided Fisher exact test between the biomarker variables and a subset of the tumor sites (breast, axillary lymph node, chest wall, and liver). Tumor sites in the Other category with 2 or more samples are: other lymph node (N=12), pleural effusion (N=8), bone (N=6), axilla (N=4), soft tissue (N=4), and lung (N=2), with all other remaining tumor sites (N=9) having only one sample per site. False discovery rates (FDR) are calculated using the P values in the biomarker status and clinical/sample characteristics comparisons of Supplemental Table S5 and S7. L-IHC, local IHC.

| **Biomarker status / tumor site** | Breast | Axillary lymph node | Chest wall | Liver | Other | Total | *P* | *FDR* |
|---|---|---|---|---|---|---|---|---|
| **HER2 L-IHC (N=306, 110 patients)** | | | | | | | | |
| 0 | 110 (57.0%) | 19 (9.8%) | 18 (9.3%) | 18 (9.3%) | 28 (14.5%) | 193 (100.0%) | 1.545E-04 | 4.326E-03 |
| Low | 84 (74.3%) | 14 (12.4%) | 2 (1.8%) | 1 (0.9%) | 12 (10.6%) | 113 (100.0%) | | |
| Total | 194 (63.4%) | 33 (10.8%) | 20 (6.5%) | 19 (6.2%) | 40 (13.1%) | 306 (100.0%) | | |
| **TROP2 status (N=161, 92 patients)** | | | | | | | | |
| Low | 9 (60.0%) | 2 (13.3%) | 0 (0.0%) | 3 (20.0%) | 1 (6.7%) | 15 (100.0%) | 0.191 | 0.667 |
| Medium/High | 95 (65.1%) | 23 (15.8%) | 11 (7.5%) | 8 (5.5%) | 9 (6.2%) | 146 (100.0%) | | |
| Total | 104 (64.6%) | 25 (15.5%) | 11 (6.8%) | 11 (6.8%) | 10 (6.2%) | 161 (100.0%) | | |
| **PD-L1 status (N=120, 80 patients)** | | | | | | | | |
| Low | 36 (56.2%) | 12 (18.8%) | 3 (4.7%) | 5 (7.8%) | 8 (12.5%) | 64 (100.0%) | 0.502 | 0.890 |
| High | 42 (75.0%) | 8 (14.3%) | 1 (1.8%) | 4 (7.1%) | 1 (1.8%) | 56 (100.0%) | | |
| Total | 78 (65.0%) | 20 (16.7%) | 4 (3.3%) | 9 (7.5%) | 9 (7.5%) | 120 (100.0%) | | |
| **sTILs status (N=184, 97 patients)** | | | | | | | | |
| Low | 26 (53.1%) | 9 (18.4%) | 4 (8.2%) | 5 (10.2%) | 5 (10.2%) | 49 (100.0%) | 0.146 | 0.667 |
| Medium/High | 95 (70.4%) | 19 (14.1%) | 4 (3.0%) | 9 (6.7%) | 8 (5.9%) | 135 (100.0%) | | |
| Total | 121 (65.8%) | 28 (15.2%) | 8 (4.3%) | 14 (7.6%) | 13 (7.1%) | 184 (100.0%) | | |

**Supplemental Table S9. Correlation between subcohort and sTILs status (pathologist assessment) across time points.** One sample per patient for each time point (DX, at diagnosis; RD, residual disease; MR, metastatic/recurrence) are included. Subcohorts: Adj, adjuvant (surgery as first intervention for early TNBC); NAC, neoadjuvant (chemotherapy as first intervention for early TNBC); dnMBC (de novo metastatic TNBC). *P* values are either two-sided Fisher exact test between the biomarker and clinical groups specified.

| **sTILs status vs Subcohort** | | | | |
|---|---|---|---|---|
| **Subcohort/sTILs status (N=145, 93 patients)** | Low | Medium/High | Total | *P* |
| Adj | 9 (34.6%) | 17 (65.4%) | 26 (100.0%) | 0.045 |
| NAC | 20 (18.2%) | 90 (81.8%) | 110 (100.0%) | |
| dnMBC | 4 (44.4%) | 5 (55.6%) | 9 (100.0%) | |
| Total | 33 (22.8%) | 112 (77.2%) | 145 (100.0%) | |
| **Subcohort/sTILs status DX (N=64, 64 patients)** | Low | Medium/High | Total | *P* |
| Adj | 3 (25.0%) | 9 (75.0%) | 12 (100.0%) | 0.191 |
| NAC | 4 (8.5%) | 43 (91.5%) | 47 (100.0%) | |
| dnMBC | 1 (20.0%) | 4 (80.0%) | 5 (100.0%) | |
| Total | 8 (12.5%) | 56 (87.5%) | 64 (100.0%) | |
| **Subcohort/sTILs status RD (N=48, 48 patients)** | Low | Medium/High | Total | *P* |
| Adj | 0 (-) | 0 (-) | 0 (100.0%) | 1.000 |
| NAC | 9 (18.8%) | 39 (81.3%) | 48 (100.0%) | |
| dnMBC | 0 (-) | 0 (-) | 0 (100.0%) | |
| Total | 9 (18.8%) | 39 (81.3%) | 48 (100.0%) | |
| **Subcohort/sTILs status DX/RD (N=82, 82 patients)** | Low | Medium/High | Total | *P* |
| Adj | 3 (25.0%) | 9 (75.0%) | 12 (100.0%) | 0.386 |
| NAC | 8 (12.3%) | 57 (87.7%) | 65 (100.0%) | |
| dnMBC | 1 (20.0%) | 4 (80.0%) | 5 (100.0%) | |
| Total | 12 (14.6%) | 70 (85.4%) | 82 (100.0%) | |
| **Subcohort/sTILs status MR (N=33, 33 patients)** | Low | Medium/High | Total | *P* |
| Adj | 6 (42.9%) | 8 (57.1%) | 14 (100.0%) | 0.634 |
| NAC | 7 (46.7%) | 8 (53.3%) | 15 (100.0%) | |
| dnMBC | 3 (75.0%) | 1 (25.0%) | 4 (100.0%) | |
| Total | 16 (48.5%) | 17 (51.5%) | 33 (100.0%) | |

**Supplemental Table S10. Correlation between age and sTILs and PD-L1 status (pathologist assessment) across time points.** One sample per patient for each time point (DX, at diagnosis; RD, residual disease; MR, metastatic/recurrence) are included. *P* values are a two-sided Wilcoxon rank sum test between the biomarker and clinical groups specified.

| **PD-L1 status vs Age** | | | | |
|---|---|---|---|---|
| **Age/PD-L1 status (N=105, 80 patients)** | Low | High | Total | *P* |
| <40 | 8 (38.1%) | 13 (61.9%) | 21 (100.0%) | 0.099 |
| 40-60 | 36 (53.7%) | 31 (46.3%) | 67 (100.0%) | |
| >60 | 11 (64.7%) | 6 (35.3%) | 17 (100.0%) | |
| Total | 55 (52.4%) | 50 (47.6%) | 105 (100.0%) | |
| **Age/PD-L1 status DX (N=41, 41 patients)** | Low | High | Total | *P* |
| <40 | 1 (12.5%) | 7 (87.5%) | 8 (100.0%) | 0.075 |
| 40-60 | 15 (57.7%) | 11 (42.3%) | 26 (100.0%) | |
| >60 | 4 (57.1%) | 3 (42.9%) | 7 (100.0%) | |
| Total | 20 (48.8%) | 21 (51.2%) | 41 (100.0%) | |
| **Age/PD-L1 status RD (N=40, 40 patients)** | Low | High | Total | *P* |
| <40 | 4 (44.4%) | 5 (55.6%) | 9 (100.0%) | 0.632 |
| 40-60 | 12 (50.0%) | 12 (50.0%) | 24 (100.0%) | |
| >60 | 4 (57.1%) | 3 (42.9%) | 7 (100.0%) | |
| Total | 20 (50.0%) | 20 (50.0%) | 40 (100.0%) | |
| **Age/PD-L1 status MR (N=24, 24 patients)** | Low | High | Total | *P* |
| <40 | 3 (75.0%) | 1 (25.0%) | 4 (100.0%) | 0.682 |
| 40-60 | 9 (52.9%) | 8 (47.1%) | 17 (100.0%) | |
| >60 | 3 (100.0%) | 0 (0.0%) | 3 (100.0%) | |
| Total | 15 (62.5%) | 9 (37.5%) | 24 (100.0%) | |
| **sTILs status vs Age** | | | | |
| **Age/sTILs status (N=152, 97 patients)** | Low | Medium/High | Total | *P* |
| <40 | 6 (17.1%) | 29 (82.9%) | 35 (100.0%) | 0.038 |
| 40-60 | 20 (21.3%) | 74 (78.7%) | 94 (100.0%) | |
| >60 | 10 (43.5%) | 13 (56.5%) | 23 (100.0%) | |
| Total | 36 (23.7%) | 116 (76.3%) | 152 (100.0%) | |
| **Age/sTILs status DX (N=67, 67 patients)** | Low | Medium/High | Total | *P* |
| <40 | 2 (13.3%) | 13 (86.7%) | 15 (100.0%) | 0.798 |
| 40-60 | 4 (9.8%) | 37 (90.2%) | 41 (100.0%) | |
| >60 | 2 (18.2%) | 9 (81.8%) | 11 (100.0%) | |
| Total | 8 (11.9%) | 59 (88.1%) | 67 (100.0%) | |
| **Age/sTILs status RD (N=50, 50 patients)** | Low | Medium/High | Total | *P* |
| <40 | 2 (13.3%) | 13 (86.7%) | 15 (100.0%) | 0.183 |
| 40-60 | 5 (17.9%) | 23 (82.1%) | 28 (100.0%) | |
| >60 | 3 (42.9%) | 4 (57.1%) | 7 (100.0%) | |
| Total | 10 (20.0%) | 40 (80.0%) | 50 (100.0%) | |
| **Age/sTILs status MR (N=35, 35 patients)** | Low | Medium/High | Total | *P* |
| <40 | 2 (40.0%) | 3 (60.0%) | 5 (100.0%) | 0.064 |
| 40-60 | 11 (44.0%) | 14 (56.0%) | 25 (100.0%) | |
| >60 | 5 (100.0%) | 0 (0.0%) | 5 (100.0%) | |
| Total | 18 (51.4%) | 17 (48.6%) | 35 (100.0%) | |

**Supplemental Figure S1. Additional HER2, TROP2, and PD-L1 quantification from sequencing and pathology.**

(A) *ERBB2* expression vs *ERBB2* copy numbers (N=45 samples, 38 patients)
(B) *ERBB2* expression (N=67 samples, 62 patients) and *ERBB2* copy numbers (N=93 samples, 102 patients) vs HER2 IHC status from the Metastatic Breast Cancer Project
(C) *ERBB2* and *MYC* copy numbers in the present cohort (N=84 samples, 57 patients) and the TNBC (N=24 samples, 20 patients) or HER2+ (N=89 samples, 68 patients) patients from the Metastatic Breast Cancer Project.

RNA-seq expression is measured in upper quartile normalized log2(TPM+1).

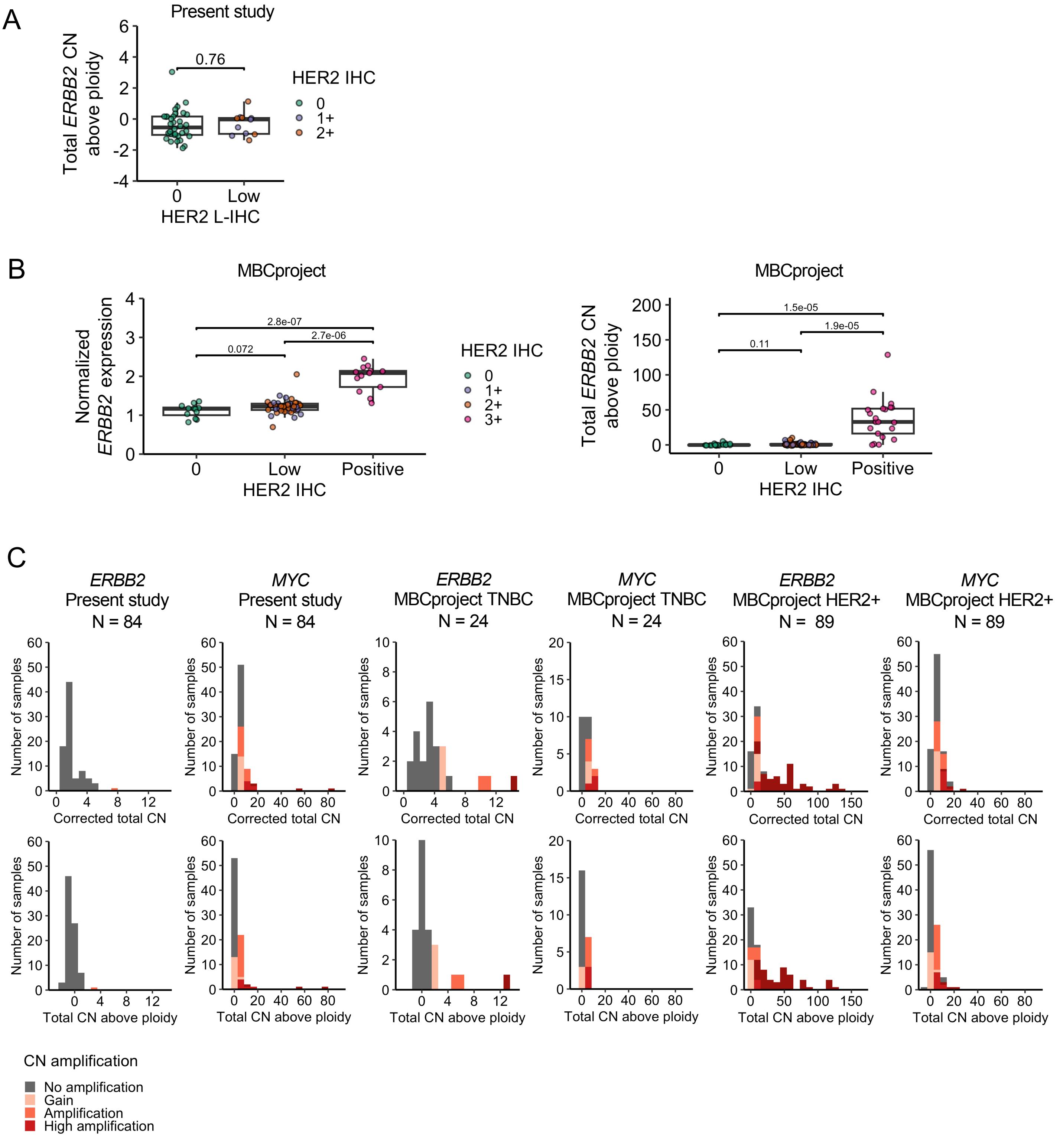

A
Present study
Total ERBB2 CN above ploidy
0.76
HER2 IHC
0
1+
2+
0
Low
HER2 L-IHC
B
MBCproject
Normalized ERBB2 expression
2.8e-07
2.7e-06
0.072
0
Low
Positive
HER2 IHC
3+
MBCproject
Total ERBB2 CN above ploidy
1.5e-05
1.9e-05
0.11
C
ERBB2
Present study
N = 84
MYC
Present study
N = 84
ERBB2
MBCproject TNBC
N = 24
MYC
MBCproject TNBC
N = 24
ERBB2
MBCproject HER2+
N = 89
MYC
MBCproject HER2+
N = 89
Number of samples
Corrected total CN
Total CN above ploidy
CN amplification
No amplification
Gain
Amplification
High amplification

**Supplemental Figure S2. Mass spectrometry protein quantification of biomarkers and correlation with pathologist-assessed biomarker quantification.**

(A) Local (L-IHC) and central (C-IHC) HER2 IHC status vs HER2 (*ERBB2)* protein expression from mass spectrometry. HER2-0 L-IHC, N=16 samples; HER2-low L-IHC, N=14 samples; C-IHC HER2-0 N=37 samples; C-IHC HER2-low N=6 samples.

(B) TROP2 pathologist-assessed quantification and TROP2 (*TACSTD2)* protein expression from mass spectrometry (N=43 samples). TROP2 low, N=6 samples; TROP2 medium/high, N=37 samples.

(C) PD-L1 tumor area positivity from tumor and immune cells and PD-L1 (*CD274)* protein expression from mass spectrometry (N=38 samples). PD-L1 low, N=23 samples; PD-L1 high, N=15 samples.

Protein expression is measured in amol/µg. Welch's t-test (two-sided) is used for statistical comparisons between groups. Pearson correlation (two-sided) is used for statistical correlation between variables.

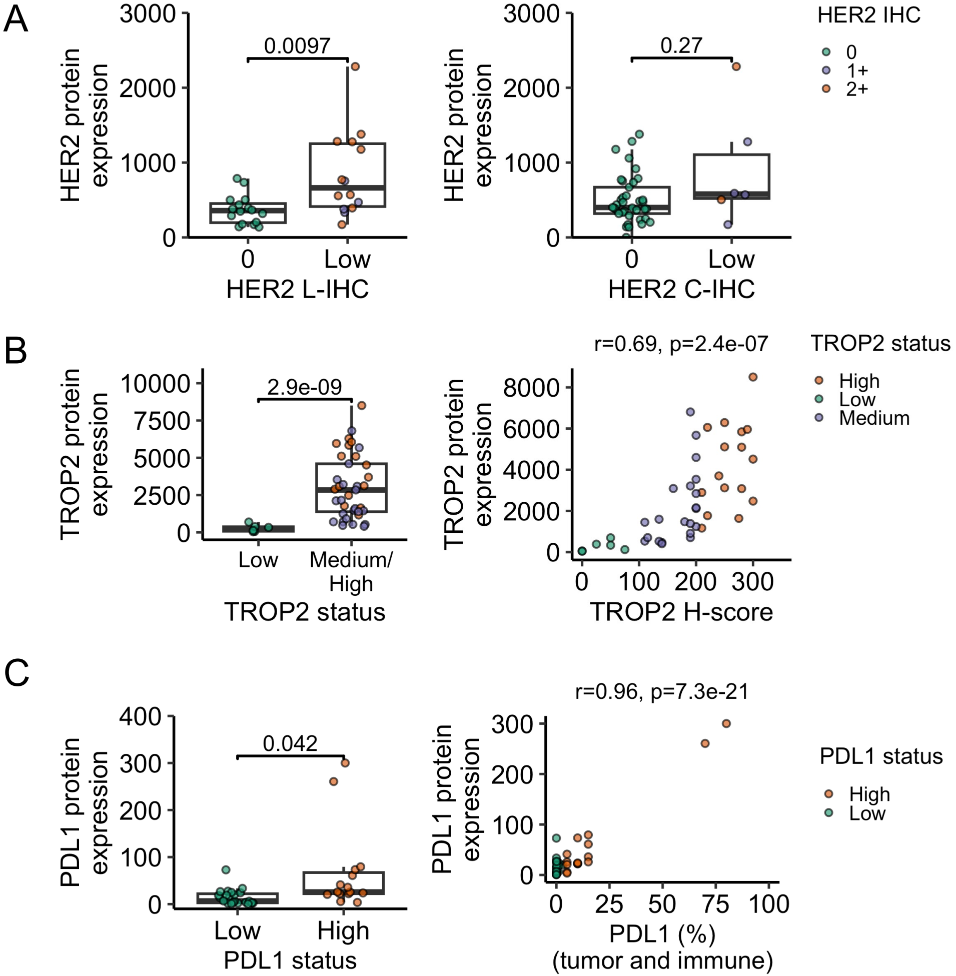
A
HER2 protein expression
3000
2000
1000
0
0.0097
0
Low
HER2 L-IHC
0.27
HER2 C-IHC
HER2 IHC
0
1+
2+
B
TROP2 protein expression
10000
7500
5000
2500
2.9e-09
Low
Medium/ High
TROP2 status
r=0.69, p=2.4e-07
8000
6000
4000
2000
100
200
300
TROP2 H-score
TROP2 status
High
Low
Medium
C
PDL1 protein expression
400
300
200
100
0.042
Low
High
PDL1 status
r=0.96, p=7.3e-21
25
50
75
100
PDL1 (%) (tumor and immune)
PDL1 status
High
Low

**Supplemental Figure S3. Computational pathology quantification of biomarkers and correlation with manual pathologist-assessed biomarker quantification.**

(A) Local (L-IHC) and central (C-IHC) HER2 IHC status from manual pathologist assessment vs HER2 QCS median membrane staining intensity (SI) from computational pathology. HER2 L-IHC 0, N=42 samples; HER2 L-IHC 1+, N=11 samples; HER2 L-IHC 2+, N=17 samples. HER2 C-IHC 0, N=60 samples; HER2 C-IHC 1+, N=17 samples; HER2 C-IHC 2+, N=9 samples

(B) TROP2 manual pathologist-assessed quantification vs TROP2 QCS median membrane SI from computational pathology (N=144 samples). TROP2 low, N=12 samples; TROP2 medium, N=49 samples; TROP2 high, N=83 samples.

(C) Stromal TILs status by manual pathologist assessment vs stromal TILs density from computational pathology (N=104 samples). sTILs low, N=16 samples; sTILs medium, N=65 samples; sTILs high, N=23 samples.

Welch’s t-test (two-sided) is used for statistical comparisons between groups.

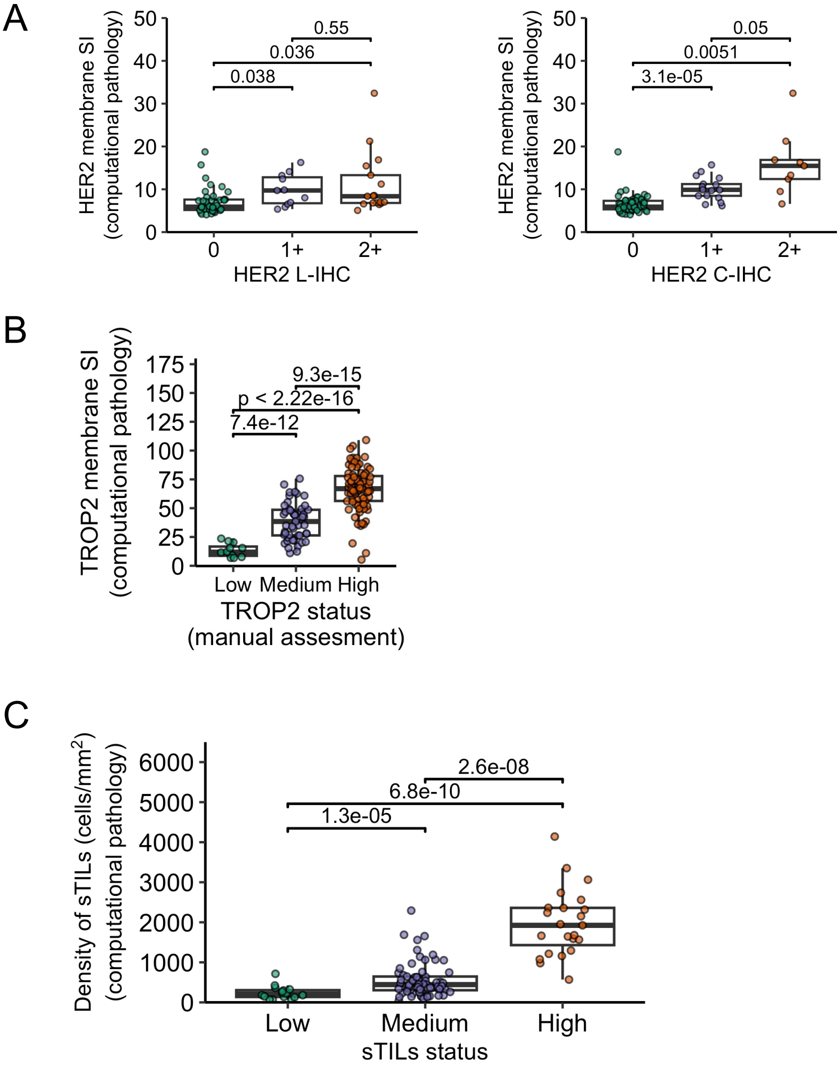
A
HER2 membrane SI (computational pathology)
50
40
30
20
10
0
0.55
0.036
0.038
0
1+
2+
HER2 L-IHC
0.05
0.0051
3.1e-05
HER2 C-IHC
B
TROP2 membrane SI (computational pathology)
175
150
125
100
75
50
25
0
9.3e-15
p < 2.22e-16
7.4e-12
Low
Medium
High
TROP2 status
(manual assesment)
C
Density of sTILs (cells/mm2) (computational pathology)
6000
5000
4000
3000
2000
1000
0
2.6e-08
6.8e-10
1.3e-05
Low
Medium
High
sTILs status
